# Inherited infertility - mapping loci associated with impaired female reproduction

**DOI:** 10.1101/2024.07.03.24309884

**Authors:** Sanni Ruotsalainen, Juha Karjalainen, Mitja Kurki, Elisa Lahtela, Matti Pirinen, Juha Riikonen, Jarmo Ritari, Silja Tammi, Jukka Partanen, Hannele Laivuori, FinnGen, Aarno Palotie, Henrike Heyne, Mark Daly, Elisabeth Widen

## Abstract

Female infertility is a common and complex health problem affecting millions of women worldwide. While multiple factors can contribute to this condition, the underlying cause remains elusive in up to 15-30% of cases. In our large genome-wide association study (GWAS) of 22,849 women with infertility and 198,989 controls from the Finnish population cohort FinnGen, we unveil a unique landscape of genetic factors associated with the disease. Our recessive analysis identified a low-frequency stop-gained mutation in *TBPL2* (p.Arg331Ter; minor allele frequency (MAF) = 1.2%) with an impact comparable to highly penetrant monogenic mutations (OR = 650, p = 4.1 ×10^-25^). While previous studies have linked the homologous gene to anovulation and sterility in knockout mice, the severe consequence of the p.Arg331Ter mutation was evidenced by homozygous carriers having significantly fewer offspring (average of 0.16) compared to women belonging to the other genotype groups (average of 1.75 offspring, p = 1.4×10^-15^). Notably, all homozygous women who had given birth had received infertility therapy. Moreover, our age-stratified analyses identified three additional genome-wide significant loci. Two loci were associated with early-onset disease (infertility diagnosed before age 30), located near *CHEK2* and within the major histocompatibility complex (MHC)-region. The third locus, associated with late-onset disease, had its lead SNP located in an intron of a lncRNA gene. Taken together, our data highlight the significance of rare recessive alleles in shaping female infertility risk. The results further provide evidence supporting specific age-dependent mechanisms underlying this complex disorder.

## INTRODUCTION

Infertility affects millions of individuals of reproductive age worldwide. WHO estimates that the lifetime prevalence of infertility in the Americas and Europe is 20% and 16.5%, respectively.^1^ Although infertility may stem from factors related both to women and men, it is commonly attributed to female factors, either entirely or in part.^2^ Ovarian dysfunction is a prevalent underlying reason, and age-related infertility is also on the rise due to delayed childbirth. Yet in 15-30% of cases, the cause of infertility remains unexplained despite thorough clinical investigation.^2,3^

Certain pre-existing conditions, such as polycystic ovarian syndrome (PCOS), endometriosis, and uterine fibroids, are associated with increased infertility risk in women. Genome-wide association studies (GWAS) have identified 20, 40, and 30 common variant loci linked to these conditions, respectively, implicating hormone signaling and cell growth pathways.^4–6^ A more recent GWAS meta-analysis of 40,024 women affected by infertility from 6 different cohorts further identified significant genetic correlations between all-cause female infertility and endometriosis and fibroids, and a correlation between anovulatory infertility and polycystic ovary syndrome (PCOS) ^7^. In that study, the majority of the 19 reported infertility loci were also associated with endometriosis, PCOS, or fibroids.^7^

While the previous GWAS analyses of infertility and its related disorders in women have targeted common genetic variation, it can be expected that rare or low-frequency recessive variants may be particularly significant causes of impaired reproduction. In fact, our previous exploration of over 40,000 coding variants across >2,000 disease phenotypes in the Finnish population cohort FinnGen, revealed a recessive association of female infertility with low-frequency variant near *PKHDL1*/*EBAG9*.^8^ That study comprised 7,980 women affected by infertility. We now present results from an expanded recessive association analysis encompassing 22,849 women affected by infertility and 198,989 controls from the FinnGen cohort. Genetic drift and historical bottlenecks have shaped the Finnish population structure leading to an enrichment of genetic variants in the 0.5-5% allele frequency range in the current population.^9^ Thus, we expect the FinnGen cohort to be highly potent and well-suited for the successful identification of such mutations impacting reproduction.

## SUBJECTS AND METHODS

### Study cohort and data

We studied female infertility in the large Finnish research project FinnGen^9^ (https://www.finngen.fi/en) launched in 2017. FinnGen is a public-private research project, combining genome and digital healthcare data on over 500,000 Finns. The nationwide research project aims to provide novel medically and therapeutically relevant insight into human diseases. FinnGen is a pre-competitive partnership of Finnish biobanks and their background organizations (universities and university hospitals) and international pharmaceutical industry partners and Finnish biobank cooperative (FINBB). All FinnGen partners are listed here: https://www.finngen.fi/en/partners. This study is based on FinnGen data release 12 including 520,210 participants of which 293,373 (56,39 %) are females.

### Definition of female infertility

The clinical disease endpoint ‘female infertility’ was constructed based on the register codes using the Finnish version of the International Classification of Diseases, 10th revision (ICD-10) diagnosis codes and harmonizing them with definitions from ICD-8 and ICD-9 as follow: at least one record of female infertility diagnosis (N97* or N98* [ICD-10], or 628* [ICD8 and 9]), or at least one record of purchased medication used for infertility treatments (ATC codes G03GA* or G03GB*) or a record of a procedure used for treatment of infertility (e.g. insemination), (NOMESCO codes: TLW00, TLW10, TLW11, TLW12, TLW14 or TLW20). Women with an infertility record including only ICD10 code N97.4 (“Female infertility associated with male factors”) were excluded from the analyses. As controls, we used women who have given birth based on data from the Medical Birth Register and the Finnish Population Register. Using these criteria, we identified 22,849 women affected by infertility and 198,989 controls.

The basic clinical characteristics of cases and controls are presented in **Table 1**. Given that the Drug Reimbursement Register contains data starting from 1995, women who have received in vitro fertilization (IVF) therapy prior to this date cannot be identified as cases unless they have received a record of infertility in the Hospital Discharge Register (HILMO). In addition, given that our resolution to distinguish between cases and controls is much higher for women suffering from reduced fertility starting from the mid-1990’s, the FinnGen women identified as cases are younger than the controls (difference of the mean = 11.99 years, p < 1× 10^-324^; **Supplementary** Figure 1). In the analyses, this age difference has been considered by using birth year as a covariate in the GWAS analyses, and when calculating the disease prevalence. In the analyses of disease enrichment, both cases and controls have been matched by birth year.

**Table 1:** Basic characteristics of the study cohort.

|  | **All** | **Cases** | **Controls** |
| --- | --- | --- | --- |
| **N (%)** | 221,838 [100] | 22,849 [10.29] | 198,989 [89.70] |
| **Age (mean [SD])<sup>a</sup>** | 61.40 [16.33] | 51.09 [12.91] | 62.59 [16.26] |
| **BMI (mean [SD])<sup>b</sup>** | 27.50 [5.72] | 27.21 [5.96] | 27.53 [5.69] |
| **Given birth (N [%])** | 216,469 [97.57] | 17,480 [76.50] | 198,989 [100] |
| **Age at first birth (mean [SD])<sup>c</sup>** | 26.20 [5.11] | 30.22 [5.77] | 25.84 [4.89] |
| **N offspring (mean [SD])** | 2.22 [1.22] | 1.50 [1.19] | 2.30 [1.19] |
| **Endometriosis (N [%])** | 16,618 [7.49] | 4,607 [20.16] | 12,011 [6.03] |
| **PCOS (N [%])** | 1,691 [0.76] | 1,011 [4.42] | 590 [0.29] |
| **Leiomyoma of uterus (N [%])** | 35 199 [15.86] | 3 346 [14.64] | 31 853 [16.01] |

<sup>a</sup> Age at end of follow-up or death
<sup>b</sup> BMI information available only for 150,636 [67.90 %] women (15,453 [67.63 %] cases and 135 183 [67.93 %] controls
<sup>c</sup> Information available only for women who have given birth at least once (n = 216,469, 17,489 cases and 198,989 controls).

5,701 (24.95 %) of cases were secondary infertility cases, i.e., they received the infertility diagnosis after having at least one child. On average, the cases with infertility who have at least one child were on average 4.37 years older than controls when giving birth for the first time (p < 1× 10^-324^; **Supplementary** Figure 2)

### Genotyping and imputation

FinnGen samples were genotyped with multiple Illumina and Affymetrix arrays (Thermo Fisher Scientific, Santa Clara, CA, USA). Genotype calls were made with GenCall and zCall algorithms for Illumina and AxiomGT1 algorithm for Affymetrix chip genotyping data batch-wise. Genotyping data produced with previous chip platforms were lifted over to build version 38 (GRCh38/hg38) following the protocol described here: dx.doi.org/10.17504/protocols.io.nqtddwn. Samples with sex discrepancies, high genotype missingness (>□2%), excess heterozygosity in common variants (allele frequency > 0.05) (±3SD) per batch, and non-Finnish ancestry were removed. Variants with high missingness (>□2%), deviation from Hardy–Weinberg equilibrium (P□<□1×10^-6^) and low minor allele count (MAC□<□3) were removed.

Pre-phasing of genotyped data was performed with Eagle 2.3.5 (https://data.broadinstitute.org/alkesgroup/Eagle/) with the default parameters, except the number of conditioning haplotypes was set to 20,000. Imputation of the genotypes was carried out by using the population-specific Sequencing Initiative Suomi (SISu) v4.2 imputation reference panel with Beagle 4.1 (version 08Jun17.d8b, https://faculty.washington.edu/browning/beagle/b4_1.html) as described in the following protocol: dx.doi.org/10.17504/protocols.io.nmndc5e. SISu v4.2 imputation reference panel was developed using the high-coverage (25–30x) whole-genome sequencing data generated at the Broad Institute of MIT and Harvard and at the McDonnell Genome Institute at Washington University, USA; and jointly processed at the Broad Institute. Variant callset was produced with Genomic Analysis Toolkit (GATK) HaplotypeCaller algorithm by following GATK best practices for variant calling. Genotype-, sample-and variant-wise quality control was applied in an iterative manner by using the Hail framework v0.2. The resulting high-quality WGS data for 3,775 individuals were phased with Eagle 2.3.5 as described above. As a post-imputation quality control, variants with INFO score < 0.7 were excluded.

### Genome-wide association study

A total of 221,838 (22,849 cases and 198,989 controls) female samples from FinnGen Data Freeze 12 were analyzed using REGENIE^10^. All models were adjusted for age at the end of follow-up or death, birth year, genotyping batch, and first ten principal components (PC). Both additive and recessive models were performed, using imputed genotypes (n variants = 21,296,962). Recessive analyses were performed for variants with at least 2 homozygous carriers for the minor allele (n variants = 15,421,131). All variants reaching genome-wide significance p-value threshold of 5×10^-8^ were considered genome-wide significant (GWS). All GWS regions were fine-mapped using SUSIE^11^, and independent genome-wide significant loci were determined as ‘good quality’ credible sets from those analyses.

To investigate the effect of advanced maternal age on reduced fertility, we also performed age-stratified GWAS. Based on the maternal age corresponding to the first record of female infertility, we divided our cases into ‘early-onset’ (‘infertility diagnosis before age of 30’, n = 9,185) and ‘late-onset’ (‘infertility diagnosis after age of 30’, n = 13,664) cases (**Supplementary** Figure 3). The age-stratified analyses were performed using both the additive and the recessive GWAS models, similarly as for the ‘all cases’ scenario described above, using the same set of controls.

### Genetic correlations and disease enrichment

To explore the connection of female infertility with underlying disorders of female reproductive traits, we calculated both 1) genetic correlations and 2) case enrichments with our female infertility endpoints across 4 disease endpoints created and analyzed in FinnGen: ‘Endometriosis’, ‘Endometriosis ESRM stage 3 or 4’, ‘Leiomyoma of uterus’ and ‘PCOS’. A complete list of endpoints analysed in FinnGen, and their definitions is available at https://www.finngen.fi/en/researchers/clinical-endpoints. We also calculated heritability estimates for both female infertility endpoints, as well as for all the four underlying disorders of female reproductive traits listed above. Summary statistics from the additive GWAS for female infertility endpoints were used to calculate genetic correlations. The genetic correlations and heritability estimates were calculated using the LDSC software.^12,13^

The prevalence in our female infertility cases and controls separately was calculated for the four underlying disorders of female reproductive traits, and its enrichment to cases vs. controls was calculated. P-values for the enrichments were calculated using Fisher’s exact test^14^. In these analyses, controls were matched with cases for their birth year, since the birth year distribution of the controls and cases were significantly different.

## RESULTS

### GWAS results

To elucidate the genetic predisposition to female infertility, we performed GWAS analyses on 22,849 women affected by from infertility and 198,989 women who had given birth without a history of infertility or infertility treatment. Both a recessive and a traditional additive model were used for the analyses. The recessive GWAS pinpointed two genome-wide significant loci: one nearby *PKHD1L1*, and another at *TBPL2*. The *TBPL2* locus has not been previously associated with female infertility or any disorder associated with infertility in GWAS. In contrast, *PKHD1L1* has previously been implicated in two GWAS analyses of female infertility-we have reported a recessive association between *PKHD1L1* and female infertility^8^, while Venkatesh and coworkers later reported a genome-wide significant association between the locus and female infertility using an additive model^7^.

We next ran the additive GWAS analysis in our cohort of cases and controls and identified genome-wide significant association at four distinct loci on chromosomes 1, 4, 6, and 8 near genes *WNT4*, *SULT1B1*, *ESR1*, and *PKHD1L1*, respectively. Statistical fine-mapping further identified a fifth independent significant association signal on *ESR1* locus (**Figure 1**). All these loci have previously been associated either with female infertility or with pre-existing conditions that increase the risk of female infertility, such as endometriosis, uterine fibroids, and PCOS (**Table 2**). Locuszoom plots for all significant GWS regions from both scans are available in **Supplementary** Figure 4 (loci identified by the additive model) and **Supplementary** Figure 5 (loci identified by the recessive model).

**Figure 1:**
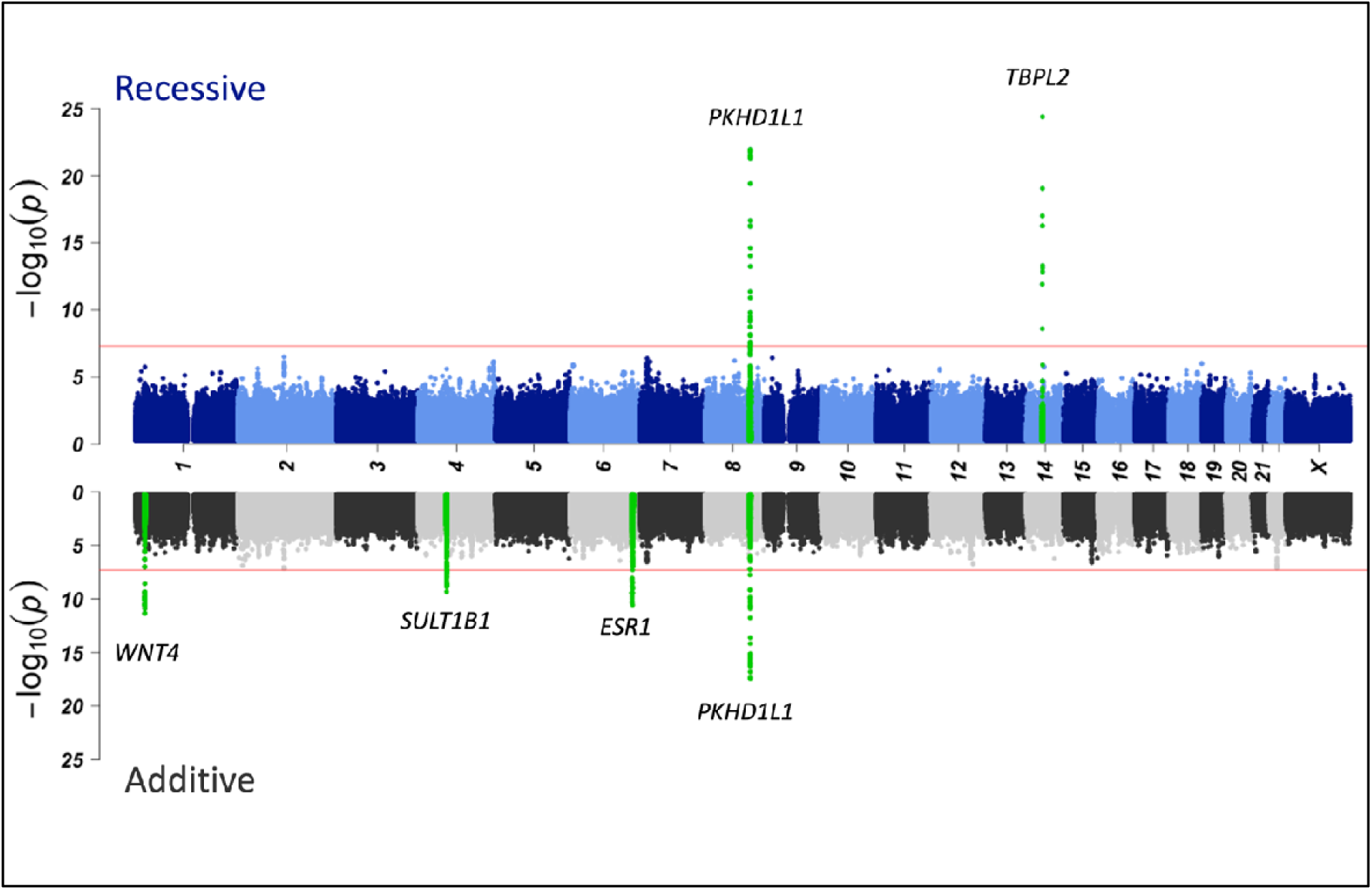
Manhattan plots for female infertility. Additive results are grey and recessive are blue dots. Genome-wide significant loci (± 500 kB) have been highlighted with green and labeled with the nearest gene of the lead variant.

**Table 2:**
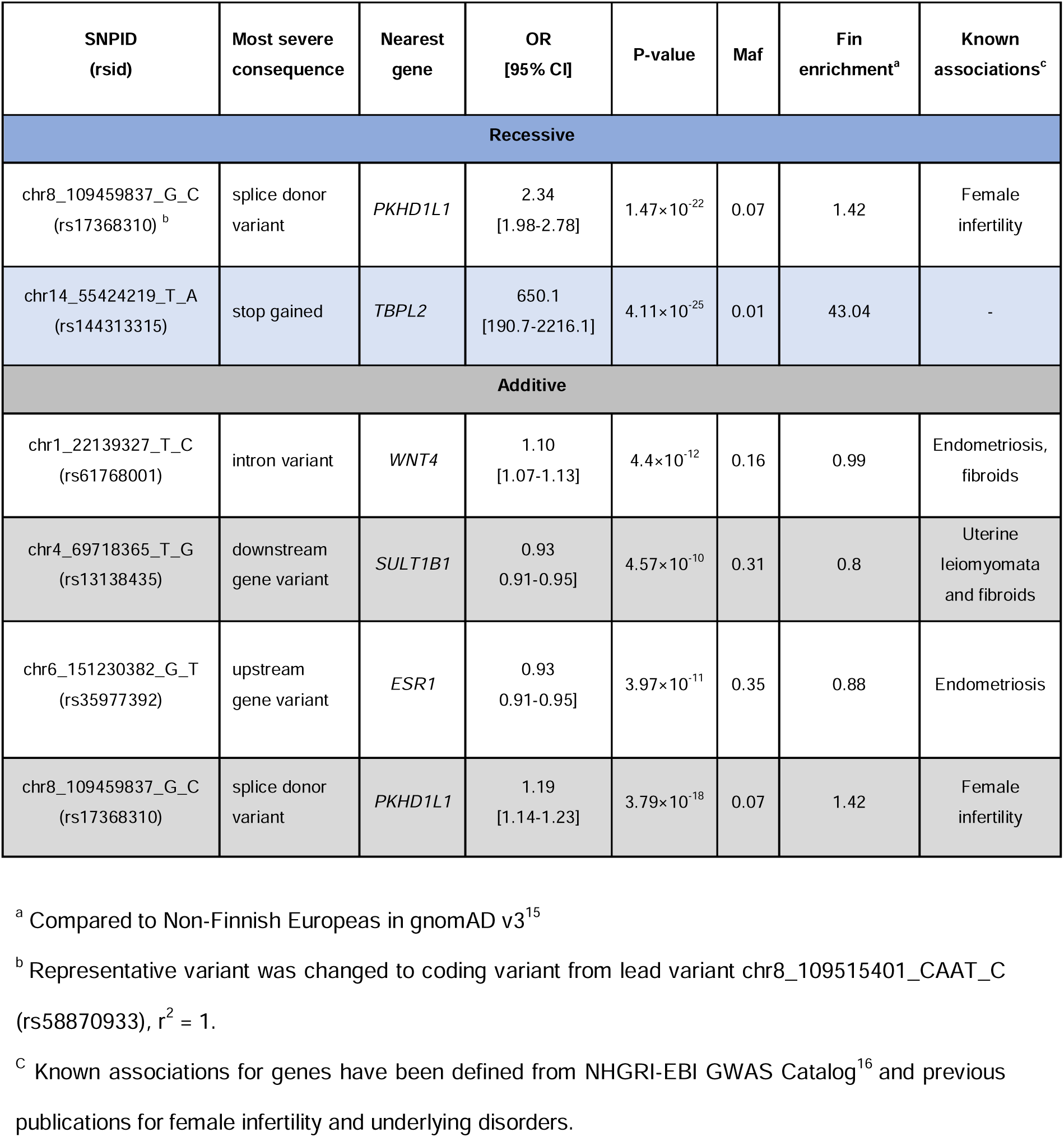
Independent GWAS hits and their representative variants identified by either additive or recessive model. The lead variant from the recessive and the additive models at the shared locus (PKHD1L1) are within the same credible set (LD = 1).

Given that the *PKHD1L1* locus yielded genome-wide significant association in both the recessive and the additive analyses, we analyzed the mode of inheritance for all identified association signals in more detail. For this purpose, we selected the lead variant (i.e. variant with the smallest p-value) at all significant loci, and plotted the proportion of affected cases in each genotype group (**Supplementary** Figure 6 and **Supplementary** Figure 7). This data confirms that both the *PKHD1L1* and *TBPL2* loci exhibit a recessive mode of action, i.e. the proportion of cases with infertility is significantly elevated only among homozygous carriers of the effect allele. In contrast, at the remaining loci, the percentage of affected cases increases across the three genotype groups consistent with an additive mode of action.

All three loci identified to have an additive effect showed evidence for pleiotropy, i.e. assessment of the infertility-associated loci by PheWas of 2,460 endpoints in FinnGen indicated association with two or more other diseases of the female reproductive tract. In contrast, the two recessive loci (near *TBPL2* and *PKHD1L1*) were specifically associated only with infertility (**Table 2**).

### *TBPL2* locus

The strongest association was observed in the recessive analysis at the *TBPL2* locus, with an OR of 650.1 (p = 4.11×10^-25^). The lead variant in this locus, rs144313315 (chr14:55424219_T/A), is a rare stop-gained mutation (p.Arg331Ter, MAF 1.1% in Finland) which is highly enriched to Finland (43 times compared to non-Finnish Europeans). The variant appears rather evenly distributed across Finland, with an allele frequency range of 0.84-2.38 % (**Supplementary** Figure 8).

Although infertility is a condition causing considerable stress to patients, most affected women will successfully conceive a child. To assess the prognosis for women carrying two copies of the *TPBL2* mutation, we analyzed the number of offspring in the entire FinnGen cohort across the *TBPL2* genotype groups. The average number of offspring among all FinnGen women with and without a history of infertility is 1.50 [SD = 1.19] vs. 1.77 [SD = 1.43], respectively. However, the average number of offspring was significantly reduced among homozygous female carriers of the p.Arg331Ter mutation. PheWAS analysis revealed a genome-wide significant recessive association between the mutation and the number of offspring (β =-1.28 SD, p = 4.25×10^-15^) with homozygous carriers having an average of only 0.16 [SD=0.58] offspring. According to the registry data, only four homozygous women for the *TBPL2* mutation had given birth, and none of them had conceived without infertility treatment **(Figure 2; Supplementary Table 1**). No such effect was observed at the *PKHD1L1* locus, where the number of offspring was only slightly reduced among homozygous carriers of the minor allele (**Supplementary** Figure 9).

**Figure 2:**
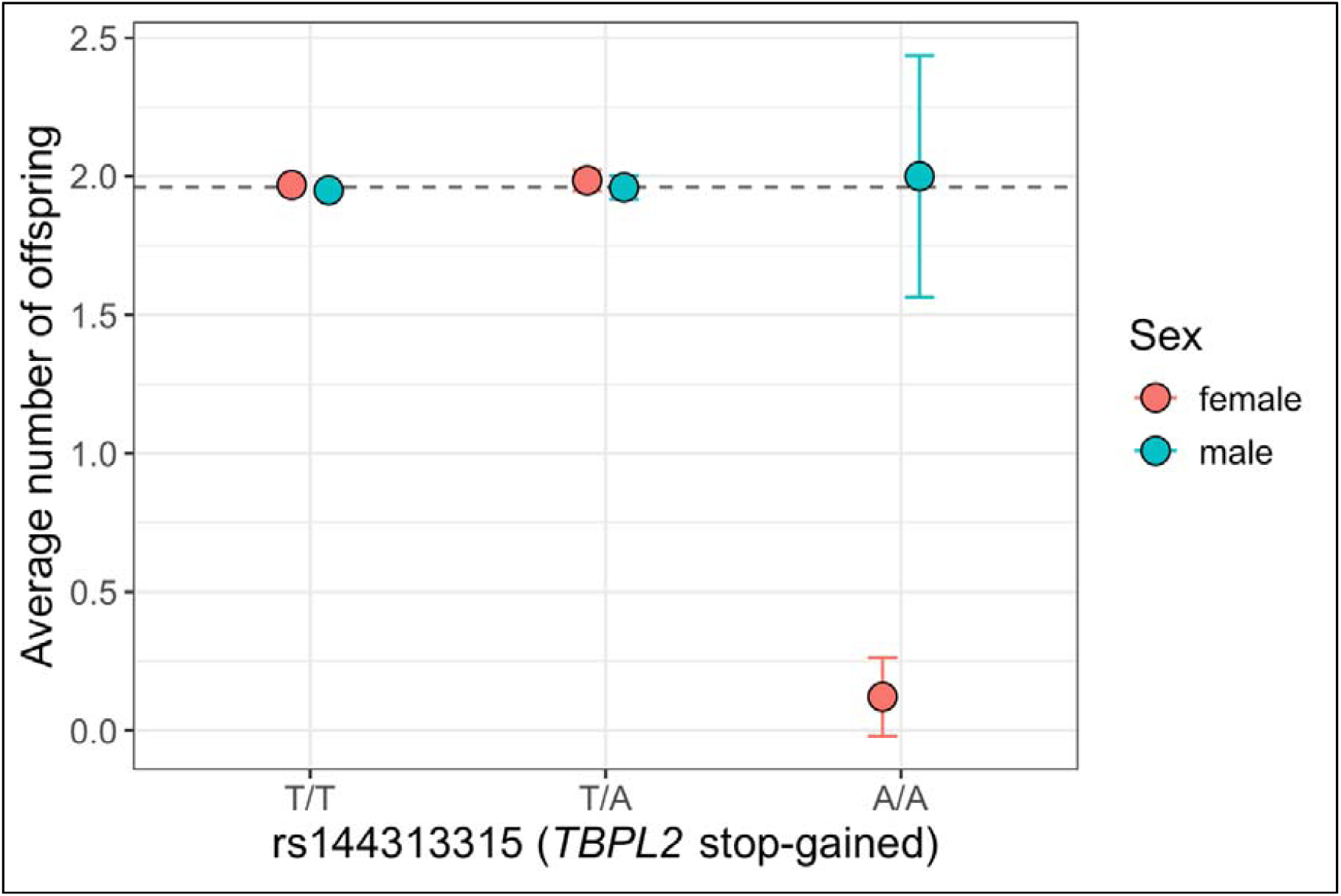
Average number of offspring for genotype groups for rs14413315 (*TBPL2* stop-gained variant) separately for females and males who have reached age 45. The dashed line represents the overall average number of offspring in FinnGen among women who have reached age 45 (=1.96).

These data indicate that the p.Arg331Ter mutation in *TPBL2* is associated with a rather severe form of infertility. These results are supported by previously published studies in mice, which reported that female mice lacking this gene are infertile female mice due to anovulation.

### Age-stratified analysis

Female fertility gradually decreases with age, starting to decline more rapidly in women in their early thirties. To assess whether genetic susceptibility varies with age, we conducted age-stratified analyses. We analyzed women diagnosed with or treated for infertility below the age of 30 (early-onset) separately from those diagnosed or treated at age 30 or above (late-onset). These age-stratified analyses identified three additional genome-wide significant loci associated with infertility: two with early-onset disease near *CHEK2* and the major histocompatibility complex (MHC) region on chromosome 9, and one with late-onset disease, with the lead SNP located in an intron of the lncRNA gene ENSG00000284418 (**Supplementary** Figure 10)

The strongest association signal exclusively associated with early-onset infertility was a frameshift mutation, c.1100delC (rs555607708), in *CHEK2*, which previously has been shown to confer increased risk for breast and ovarian cancer. The mutation is additionally associated with leiomyoma, thyroid cancer, and myeloproliferative disorders in FinnGen. A recent study including women from FinnGen and the Estonian biobank further reported that the mutation is associated with an increased risk for PCOS (OR = 13.46 [5.68–31.89], p = 1.68×10^-9^)^17^

The other association signal exclusively associated with early-onset infertility was within the major histocompatibility complex (MHC) region. To pinpoint the exact location of the signal, we imputed the classical HLA alleles as well as *MICA* and *MICB*^18^ and tested these alleles for association with our female infertility endpoints. None of the tested variants showed significant association for any of the infertility endpoint tested. The complete association results of these additional association tests across the MHC region are presented in **Supplementary Table 2**.

### Genetic correlations and case enrichments among cases

Given that three of the significantly associated loci detected by the primary GWAS analyses showed pleiotropic association with female reproductive system disorders, such as PCOS, endometriosis, and leiomyoma of the uterus, we next examined their genetic overlap with infertility on a genomic scale. The strongest genetic correlation (rg) was observed between infertility and endometriosis (rg = 0.46, p = 2.69×10^-15)^, with similar correlation estimates with endometriosis found among both early-onset and late-onset cases (**Supplementary** Figure 11). In contrast, although there was a significant enrichment of PCOS among women affected by infertility (**Table 3**), the genetic correlation between the two diseases was only 0.22 (p = 0.014). Interestingly, both the genetic correlation and case enrichment with PCOS increased among early-onset cases. Heritability estimates for all analyzed female infertility-related endpoints are presented in **Supplementary** Figure 12.

**Table 3:** Genetic correlations with female infertility (rg) and enrichment of female infertility cases (Enrichment) among four underlying disorders for female reproduction.

| Phenotype | N cases | rg [95% CI] | P-value (rg) | Enrichment | P-value (Enr) |
| --- | --- | --- | --- | --- | --- |
| Endometriosis | 20 190 | 0.46 [0.35-0.58] | $2.69 \times 10^{-15}$ | 3.83 | $< 1 \times 10^{-324}$ |
| Endometriosis ASRM stages 3,4 | 10 127 | 0.48 [0.35-0.61] | $1.76 \times 10^{-13}$ | 5.53 | $< 1 \times 10^{-324}$ |
| Leiomyoma of uterus | 42 107 | 0.30 [0.19-0.41] | $8.78 \times 10^{-08}$ | 1.44 | $1.01 \times 10^{-73}$ |
| Polycystic ovarian syndrome (PCOS) | 2 214 | 0.22 [0.04-0.39] | 0.014 | 9.27 | $< 1 \times 10^{-324}$ |

## DISCUSSION

Our GWAS analyses, encompassing more than 20,000 Finnish women affected by infertility and 198,000 controls, showcase a diverse landscape of associated loci. The analyses reveal that a portion of unexplained female infertility may be attributed to high-impact recessively acting disruptive mutations present at >1% frequency in the population. Specifically, our analyses point to a mutation in the Tata-box binding protein-like 2 gene (*TBPL2)*, which encodes a transcription factor protein crucial for transcription initiation during oocyte growth. Based on our findings, this mutation is a significant contributor to female infertility in Finland.^19^ The mutation introduces a premature stop codon (p.Arg331Ter) in the terminal DNA-binding part of the protein, and can hence be expected to significantly impair the transcription of the downstream targets. In our follow-up analyses, the *TBPL2* mutation was associated only with infertility and no other diseases. The variant’s association with a smaller number of offspring suggests that the mutation is associated with severe infertility Prior data have indeed shown that the absence of *TBPL2* leads to oocytes failing to develop and renders female mice sterile. ^20^

According to data from GnomAD (v3.1.2), loss-of-function mutations in *TBPL2* are rare, with the stop-gained mutation associated with infertility in this study being an exception. Apart from our findings, there are only scattered reports of coding mutations in *TBPL2* detected in women with infertility. Three reports describe homozygous and compound heterozygous stop-gained mutations (c.788+3A>G, p.Arg233Ter and c.802C>T, p.Arg268Ter) in women with infertility from four unrelated Chinese families.^21,22^ In a fourth report, a homozygous missense mutation was linked to infertility, oocyte maturation arrest, and degradation in two sisters from a consanguineous family.^23^ Notably, all published cases have exhibited treatment-resistant primary infertility. In our study, most homozygous mutation carriers had not given birth. However, based on data from the Medical Birth Register, two out of 41 carriers had given birth to one child, whereas two carriers had given birth to more than one child. All four women had undergone IVF therapy, although more detailed information on the potential use of donor oocytes is not available.

Over the past few decades, there has been a considerable shift in childbearing demographics in developed countries, characterized by more women choosing to delay childbearing.^24,25^ Since a woman’s age is one of the key factors influencing fertility, this trend has resulted in a higher average age at which women first experience infertility. This pattern is also evident in FinnGen, where we observe steadily increasing ages at first birth among women born between the 1950s and the 1980s (**Supplementary** Figure 13). Interestingly, previous research indicates that the causes of infertility may differ between older and younger women, with those over 35 years of age being nearly twice as likely to experience unexplained infertility compared to younger women.^26^

In support of specific age-dependent mechanisms, our GWAS analyses suggest that some associated loci indeed exert their effect in an age-dependent manner. Some loci, such as *TBPL2*, are uniformly associated with infertility in both early and late-onset cases, but we identified two loci uniquely associated with infertility only among women diagnosed before age 30, and one locus associated with late-onset disease. One of the loci associated with early-onset infertility is a frameshift alteration (c.1100delC) in the tumor suppressor gene *CHEK2,* associated with a 32% lifetime risk for breast cancer.^27^ Finding association with early-onset infertility likely reflects a link with PCOS, as the c.1100delC mutation is significantly associated with PCOS in FinnGen ^16^, and the proportion of PCOS cases is increased among women with early-onset infertility compared to all infertility cases (0.077 vs. 0.046, respectively).

The second locus associated with the early-onset infertility was the major histocompatibility complex (MHC) region on chromosome 6. The data indicate that the association is rather sharply defined to a region between 30018523 and 33018523 bp on chromosome 6, and fine-mapping analyses suggest that the signal is not associated with the classical MHC effect. The classical MHC genes encode proteins involved in antigen presentation to T cells, whereas a key function for non-classical MHC I molecules is to mediate inhibitory or activating stimuli in natural killer (NK) cells. Given that the survival of the embryo in the uterus depends on the maintenance of immune tolerance at the maternal-fetal interface, and that the pregnant uterus is predominantly populated with NK cells, one may speculate that the association findings tag an underlying mechanism that might be related to local immune tolerance.^28^ Further functional studies are nonetheless warranted to explore and verify such an effect.

Taken together, our data point towards a unique landscape of genetic factors associated with female infertility. The results specifically highlight the significant role of recessive low-frequency variants, with the stop-gained mutation in *TBPL2* exhibiting an impact comparable to highly penetrant monogenic mutations.

## Data Availability

The FinnGen data may be accessed through Finnish Biobanks' FinnBB portal (www.finbb.fi) and THL Biobank data through THL Biobank (https://thl.fi/en/web/thl-biobank).

## Ethics statement

Study subjects in FinnGen provided informed consent for biobank research, based on the Finnish Biobank Act. Alternatively, separate research cohorts, collected prior the Finnish Biobank Act came into effect (in September 2013) and start of FinnGen (August 2017), were collected based on study-specific consents and later transferred to the Finnish biobanks after approval by Fimea (Finnish Medicines Agency), the National Supervisory Authority for Welfare and Health. Recruitment protocols followed the biobank protocols approved by Fimea. The Coordinating Ethics Committee of the Hospital District of Helsinki and Uusimaa (HUS) statement number for the FinnGen study is Nr HUS/990/2017.

The FinnGen study is approved by Finnish Institute for Health and Welfare (permit numbers: THL/2031/6.02.00/2017, THL/1101/5.05.00/2017, THL/341/6.02.00/2018, THL/2222/6.02.00/2018, THL/283/6.02.00/2019, THL/1721/5.05.00/2019 and THL/1524/5.05.00/2020), Digital and population data service agency (permit numbers: VRK43431/2017-3, VRK/6909/2018-3, VRK/4415/2019-3), the Social Insurance Institution (permit numbers: KELA 58/522/2017, KELA 131/522/2018, KELA 70/522/2019, KELA 98/522/2019, KELA 134/522/2019, KELA 138/522/2019, KELA 2/522/2020, KELA 16/522/2020), Findata permit numbers

THL/2364/14.02/2020, THL/4055/14.06.00/2020, THL/3433/14.06.00/2020, THL/4432/14.06/2020, THL/5189/14.06/2020, THL/5894/14.06.00/2020, THL/6619/14.06.00/2020, THL/209/14.06.00/2021, THL/688/14.06.00/2021, THL/1284/14.06.00/2021, THL/1965/14.06.00/2021, THL/5546/14.02.00/2020, THL/2658/14.06.00/2021, THL/4235/14.06.00/2021, Statistics Finland (permit numbers: TK-53-1041-17 and TK/143/07.03.00/2020 (earlier TK-53-90-20) TK/1735/07.03.00/2021, TK/3112/07.03.00/2021) and Finnish Registry for Kidney Diseases permission/extract from the meeting minutes on 4th July 2019.

The Biobank Access Decisions for FinnGen samples and data utilized in FinnGen Data Freeze 11 include: THL Biobank BB2017_55, BB2017_111, BB2018_19, BB_2018_34, BB_2018_67, BB2018_71, BB2019_7, BB2019_8, BB2019_26, BB2020_1, BB2021_65, Finnish Red Cross Blood Service Biobank 7.12.2017, Helsinki Biobank HUS/359/2017, HUS/248/2020, HUS/430/2021 §28, §29, HUS/150/2022 §12, §13, §14, §15, §16, §17, §18, §23, §58, §59, HUS/128/2023 §18, Auria Biobank AB17-5154 and amendment #1 (August 17 2020) and amendments BB_2021-0140, BB_2021-0156 (August 26 2021, Feb 2 2022), BB_2021-0169, BB_2021-0179, BB_2021-0161, AB20-5926 and amendment #1 (April 23 2020) and it′s modifications (Sep 22 2021), BB_2022-0262, BB_2022-0256, Biobank Borealis of Northern Finland_2017_1013, 2021_5010, 2021_5010 Amendment, 2021_5018, 2021_5018 Amendment, 2021_5015, 2021_5015 Amendment, 2021_5015 Amendment_2, 2021_5023, 2021_5023 Amendment, 2021_5023 Amendment_2, 2021_5017, 2021_5017 Amendment, 2022_6001, 2022_6001 Amendment, 2022_6006 Amendment, 2022_6006 Amendment, 2022_6006 Amendment_2, BB22-0067, 2022_0262, 2022_0262 Amendment, Biobank of Eastern Finland 1186/2018 and amendment 22§/2020, 53§/2021, 13§/2022, 14§/2022, 15§/2022, 27§/2022, 28§/2022, 29§/2022, 33§/2022, 35§/2022, 36§/2022, 37§/2022, 39§/2022, 7§/2023, 32§/2023, 33§/2023, 34§/2023, 35§/2023, 36§/2023, 37§/2023, 38§/2023, 39§/2023, 40§/2023, 41§/2023, Finnish Clinical Biobank Tampere MH0004 and amendments (21.02.2020 & 06.10.2020), BB2021-0140 8§/2021, 9§/2021, §9/2022, §10/2022, §12/2022, 13§/2022, §20/2022, §21/2022, §22/2022, §23/2022, 28§/2022, 29§/2022, 30§/2022, 31§/2022, 32§/2022, 38§/2022, 40§/2022, 42§/2022, 1§/2023, Central Finland Biobank 1-2017, BB_2021-0161, BB_2021-0169, BB_2021-0179, BB_2021-0170, BB_2022-0256, BB_2022-0262, BB22-0067, Decision allowing to continue data processing until 31st Aug 2024 for projects: BB_2021-0179, BB22-0067,BB_2022-0262, BB_2021-0170, BB_2021-0164, BB_2021-0161, and BB_2021-0169, and Terveystalo Biobank STB 2018001 and amendment 25th Aug 2020, Finnish Hematological Registry and Clinical Biobank decision 18th June 2021, Arctic biobank P0844: ARC_2021_1001.

## Data and code availability

Individual level data: The Finnish biobank data can be accessed through the Fingenious® services (https://site.fingenious.fi/en/) managed by FINBB. Finnish Health register data can be applied from Findata (https://findata.fi/en/data/).

Summary statistics: Summary statistics from each data release will be made publicly available after a one year embargo period and can be accessed freely from: www.finngen.fi/en/access_results.

## Acknowledgements

We want to acknowledge the participants and investigators of the FinnGen study. The FinnGen project is funded by two grants from Business Finland (HUS 4685/31/2016 and UH 4386/31/2016) and the following industry partners: AbbVie Inc., AstraZeneca UK Ltd, Biogen MA Inc., Bristol Myers Squibb (and Celgene Corporation & Celgene International II Sàrl), Genentech Inc., Merck Sharp & Dohme LCC, Pfizer Inc., GlaxoSmithKline Intellectual Property Development Ltd., Sanofi US Services Inc., Maze Therapeutics Inc., Janssen Biotech Inc, Novartis AG, and Boehringer Ingelheim International GmbH. Following biobanks are acknowledged for delivering biobank samples to FinnGen: Auria Biobank (www.auria.fi/biopankki), THL Biobank (www.thl.fi/biobank), Helsinki Biobank (www.helsinginbiopankki.fi), Biobank Borealis of Northern Finland (https://www.ppshp.fi/Tutkimus-ja-opetus/Biopankki/Pages/Biobank-Borealis-briefly-in-English.aspx), Finnish Clinical Biobank Tampere (www.tays.fi/en-US/Research_and_development/Finnish_Clinical_Biobank_Tampere), Biobank of Eastern Finland (www.ita-suomenbiopankki.fi/en), Central Finland Biobank (www.ksshp.fi/fi-FI/Potilaalle/Biopankki), Finnish Red Cross Blood Service Biobank (www.veripalvelu.fi/verenluovutus/biopankkitoiminta), Terveystalo Biobank (www.terveystalo.com/fi/Yritystietoa/Terveystalo-Biopankki/Biopankki/) and Arctic Biobank (https://www.oulu.fi/en/university/faculties-and-units/faculty-medicine/northern-finland-birth-cohorts-and-arctic-biobank). All Finnish Biobanks are members of BBMRI.fi infrastructure (https://www.bbmri-eric.eu/national-nodes/finland/). Finnish Biobank Cooperative-FINBB (https://finbb.fi/) is the coordinator of BBMRI-ERIC operations in Finland. The Finnish biobank data can be accessed through the Fingenious® services (https://site.fingenious.fi/en/) managed by FINBB.

---

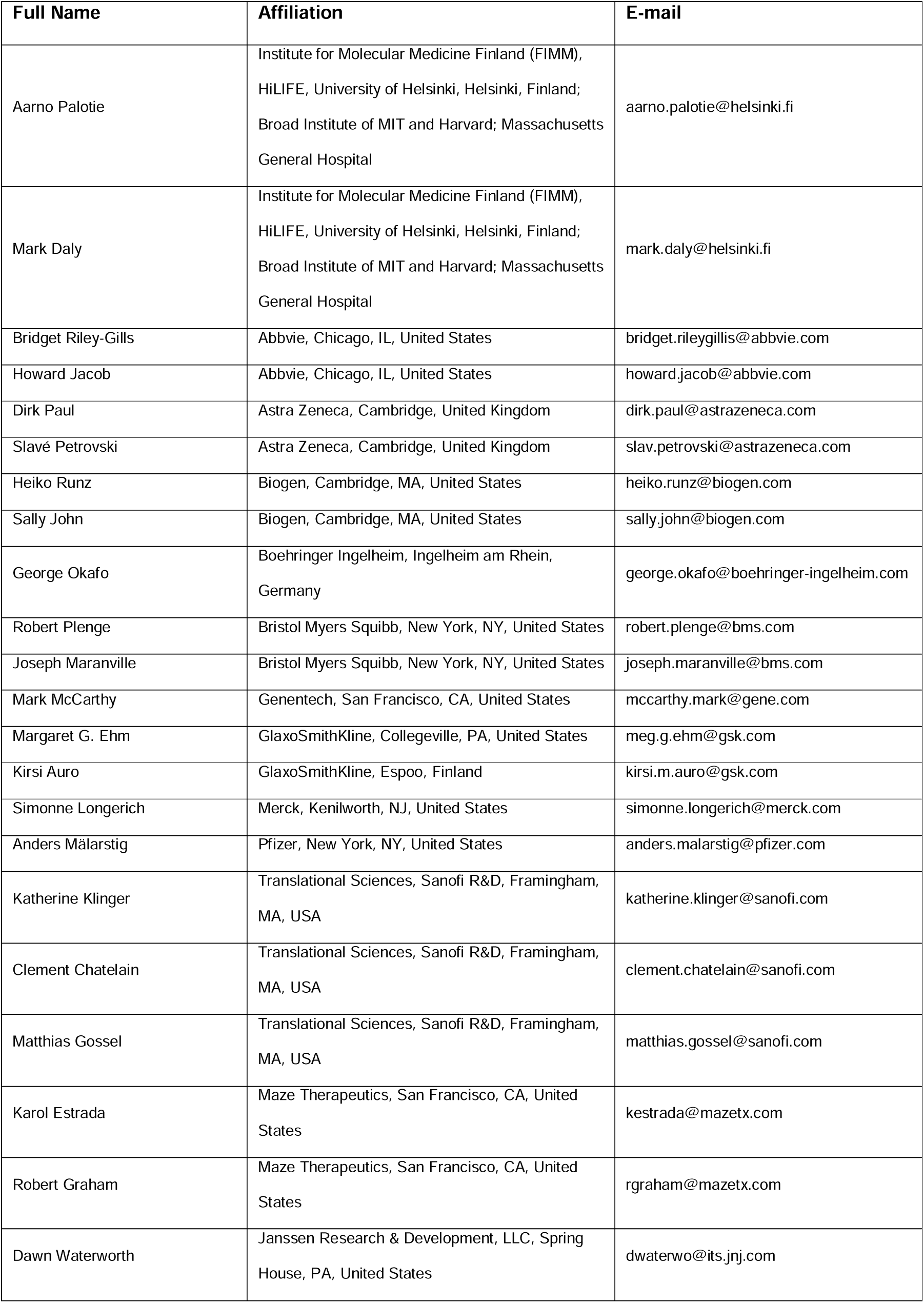
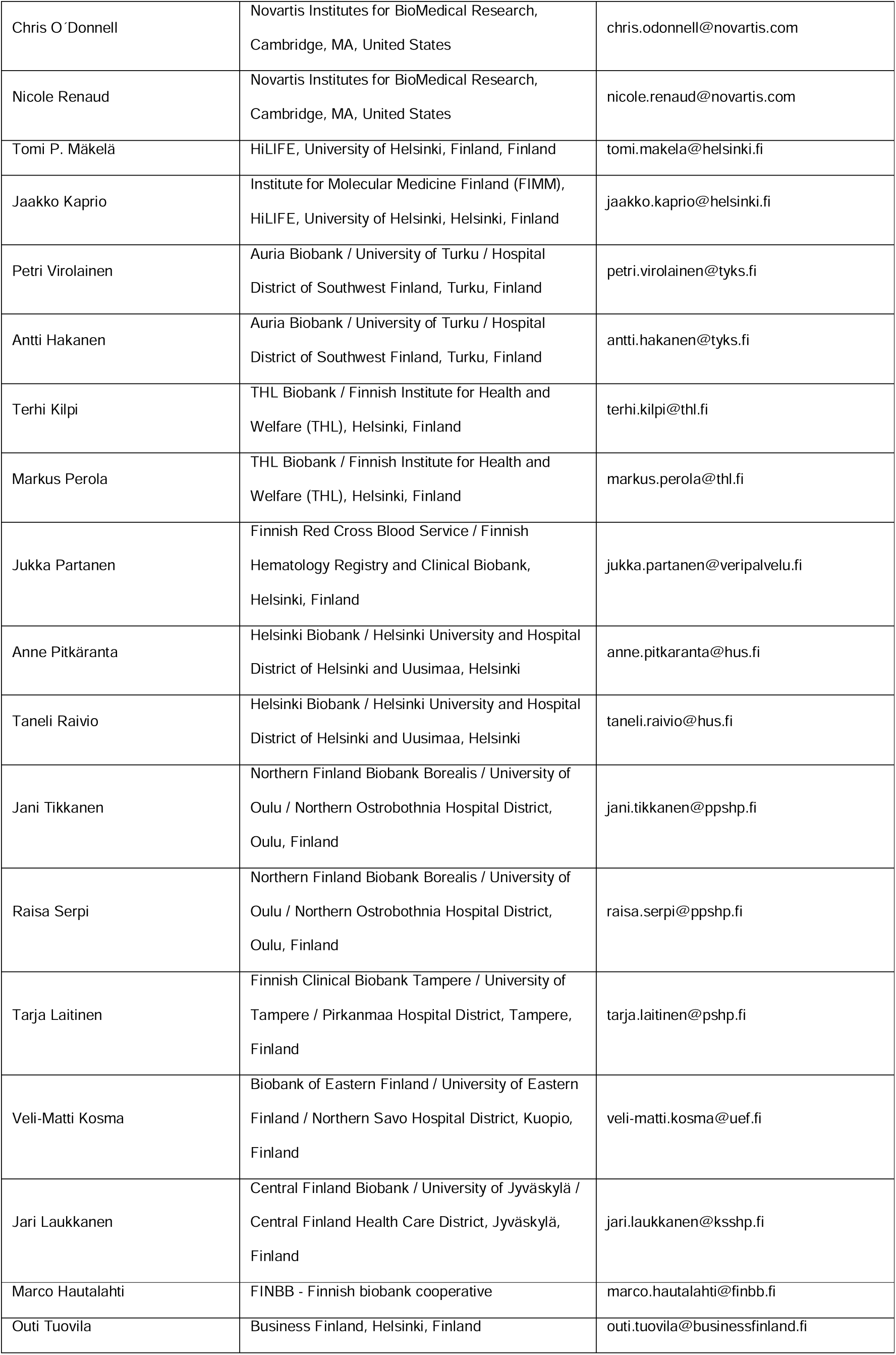
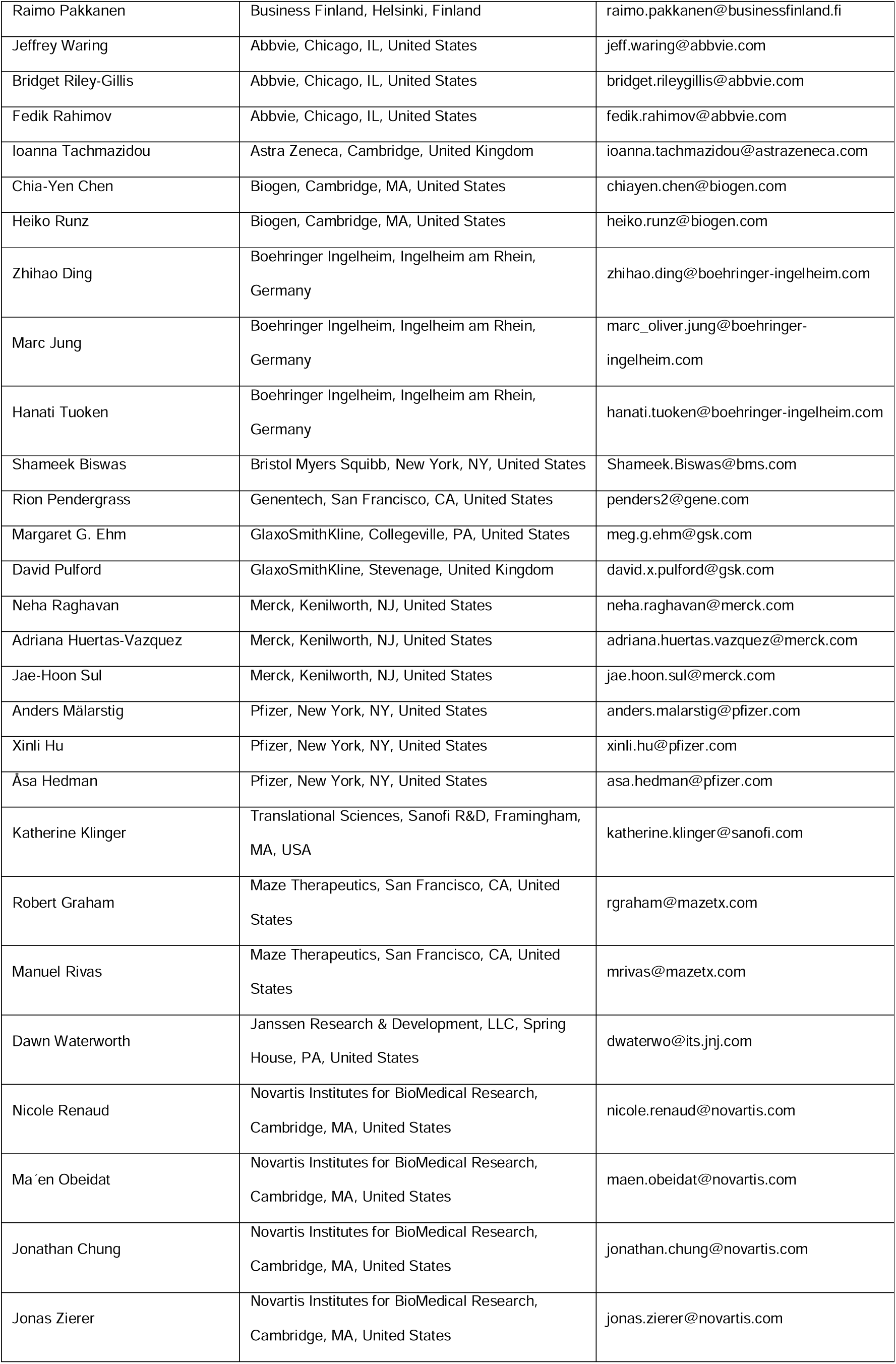
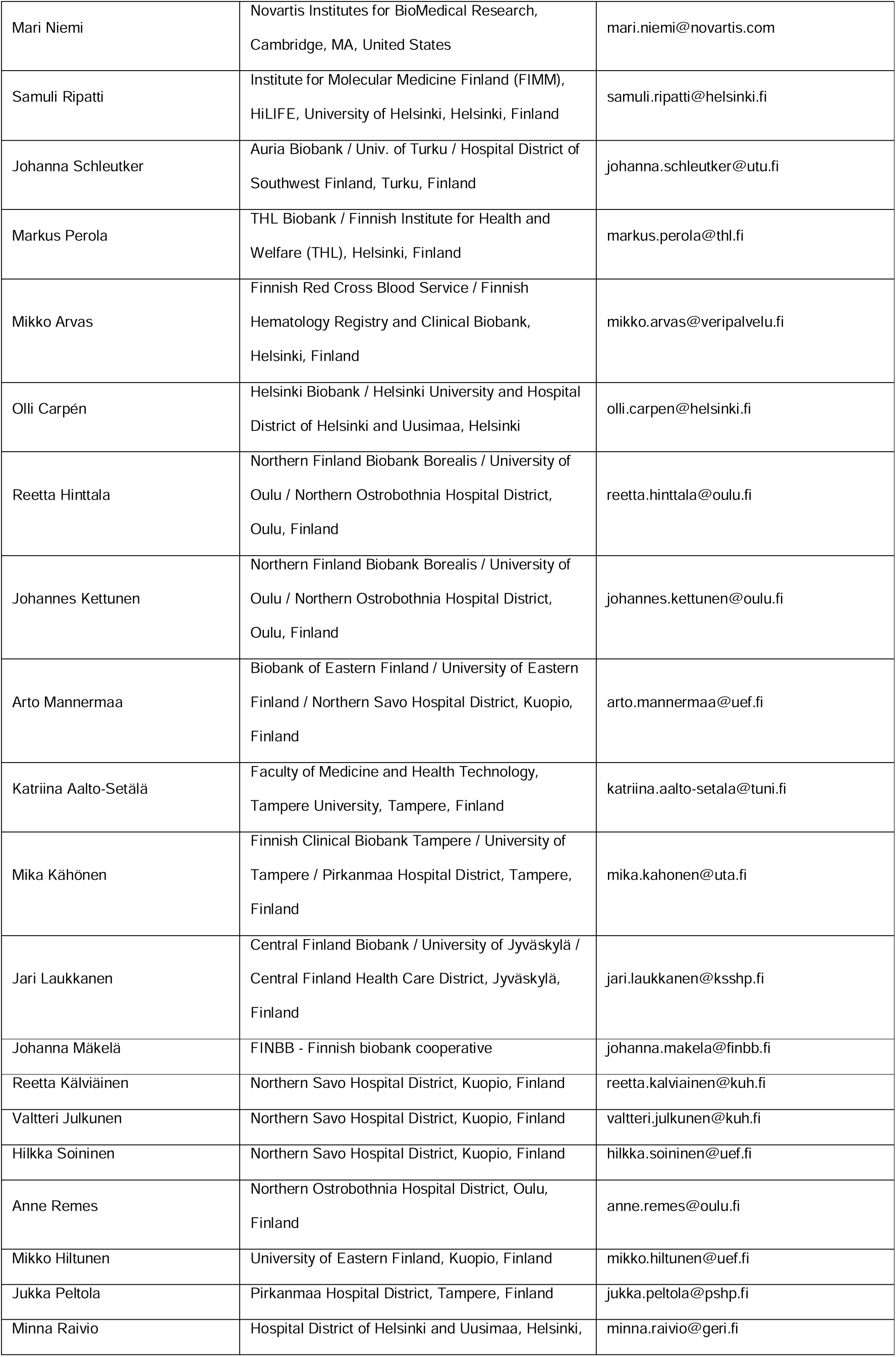
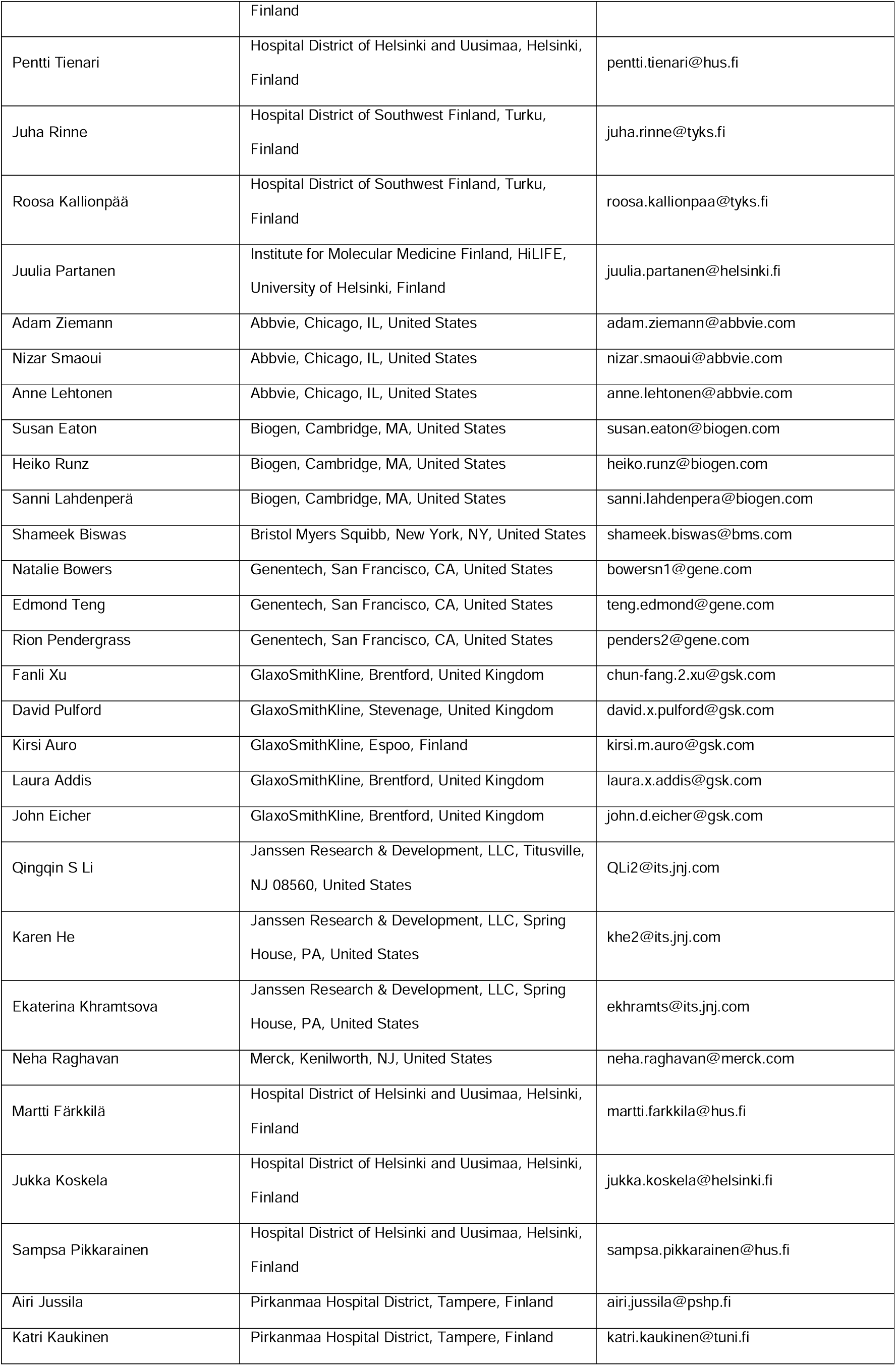
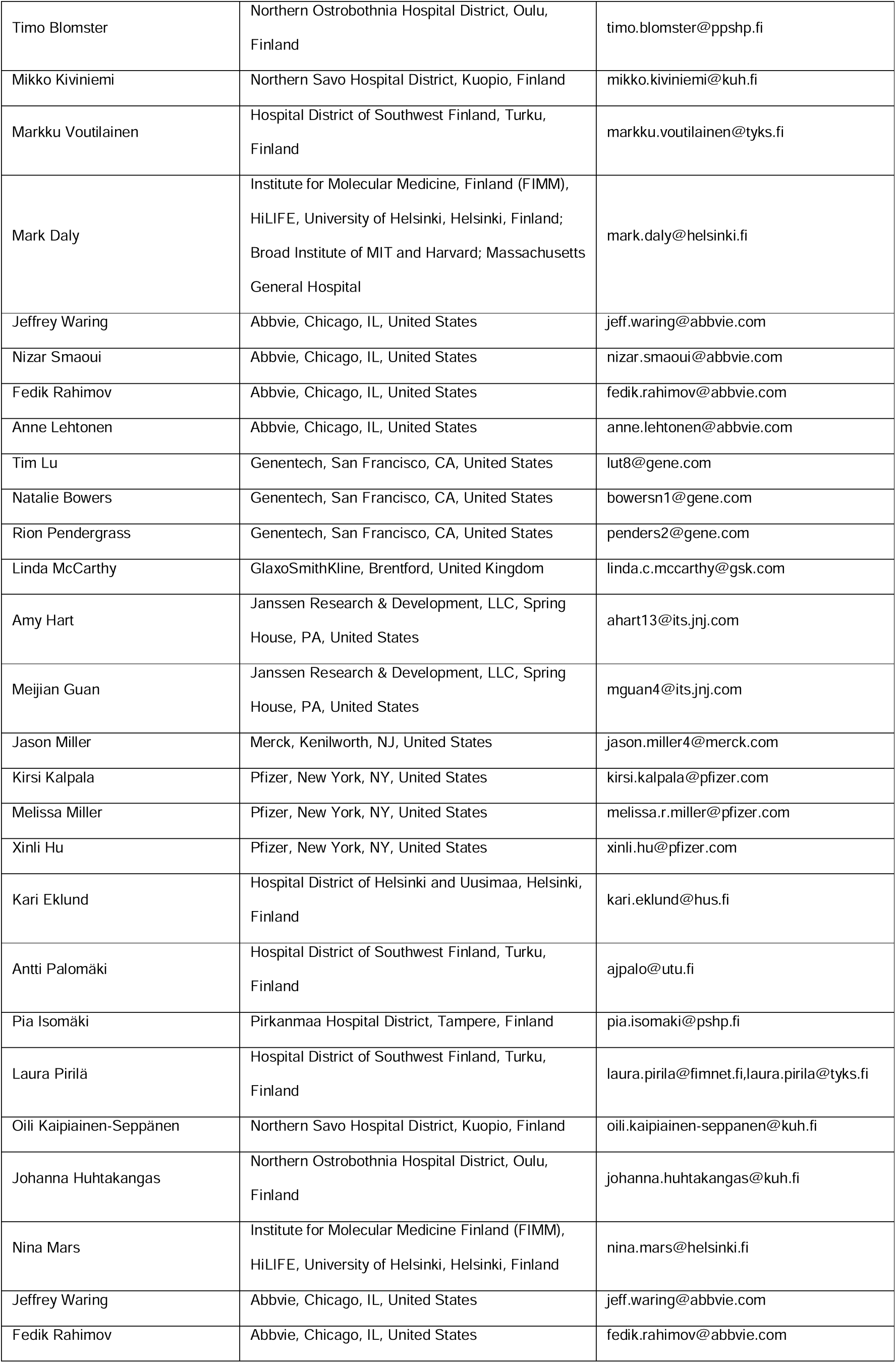
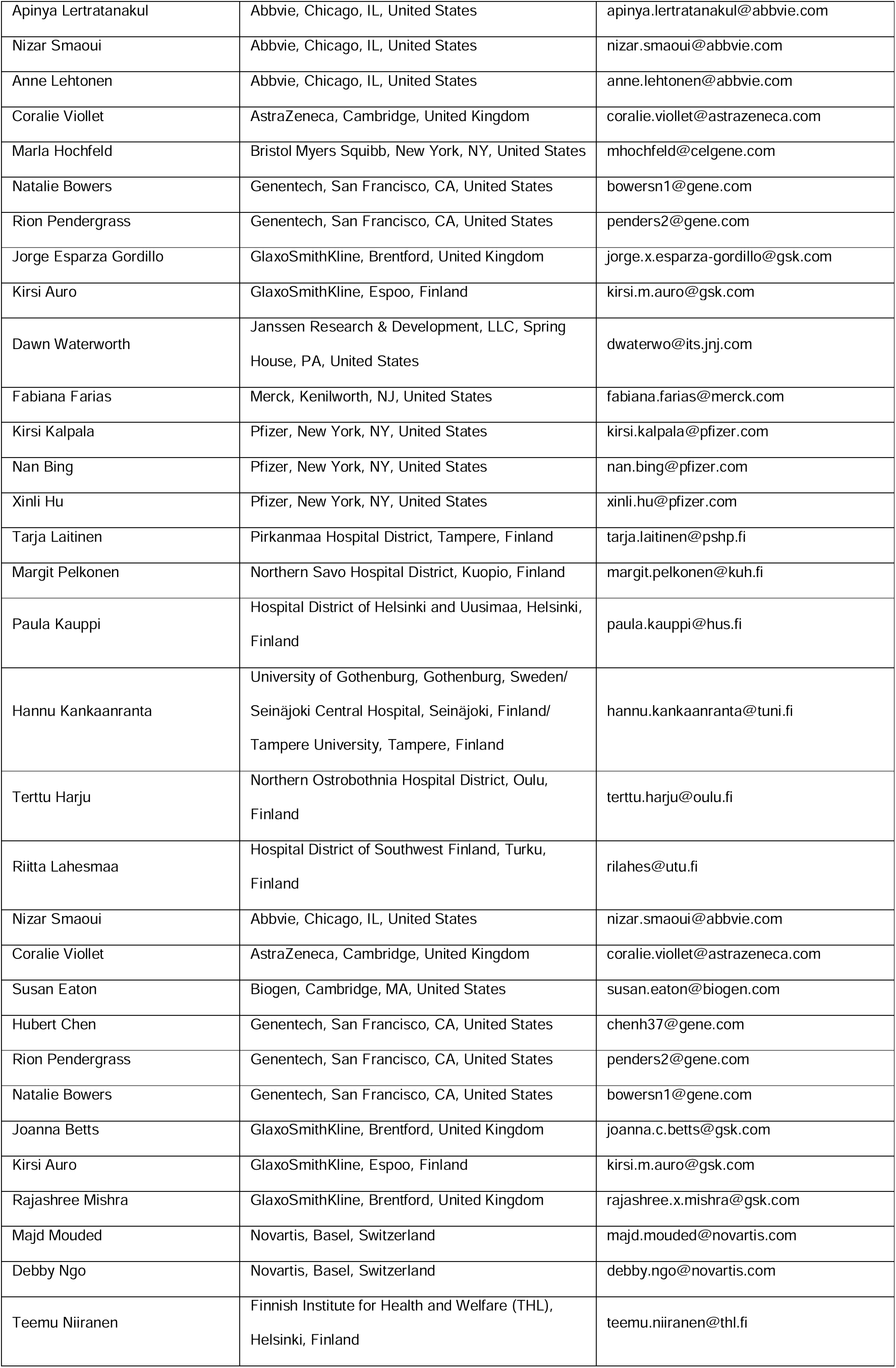
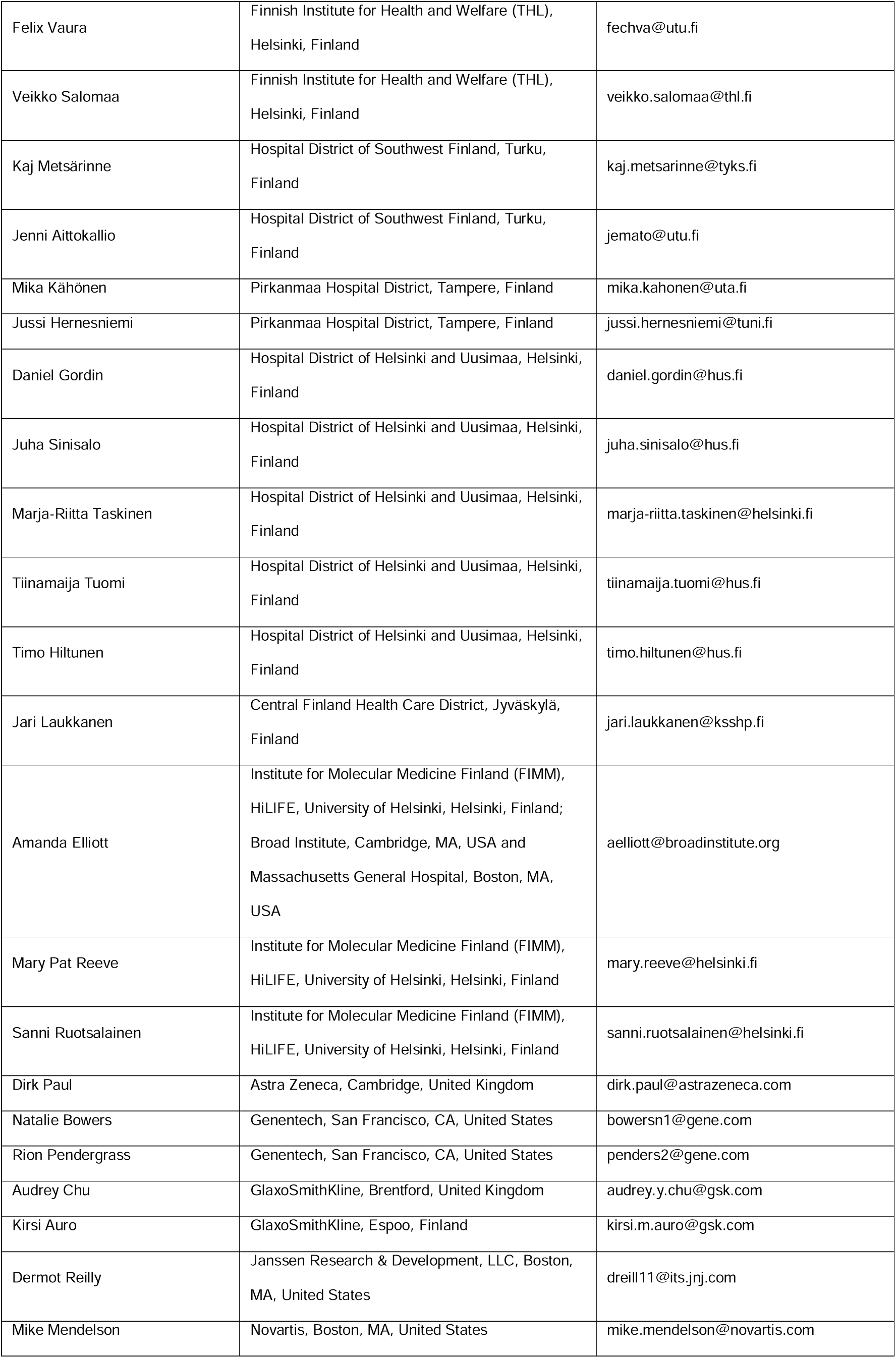
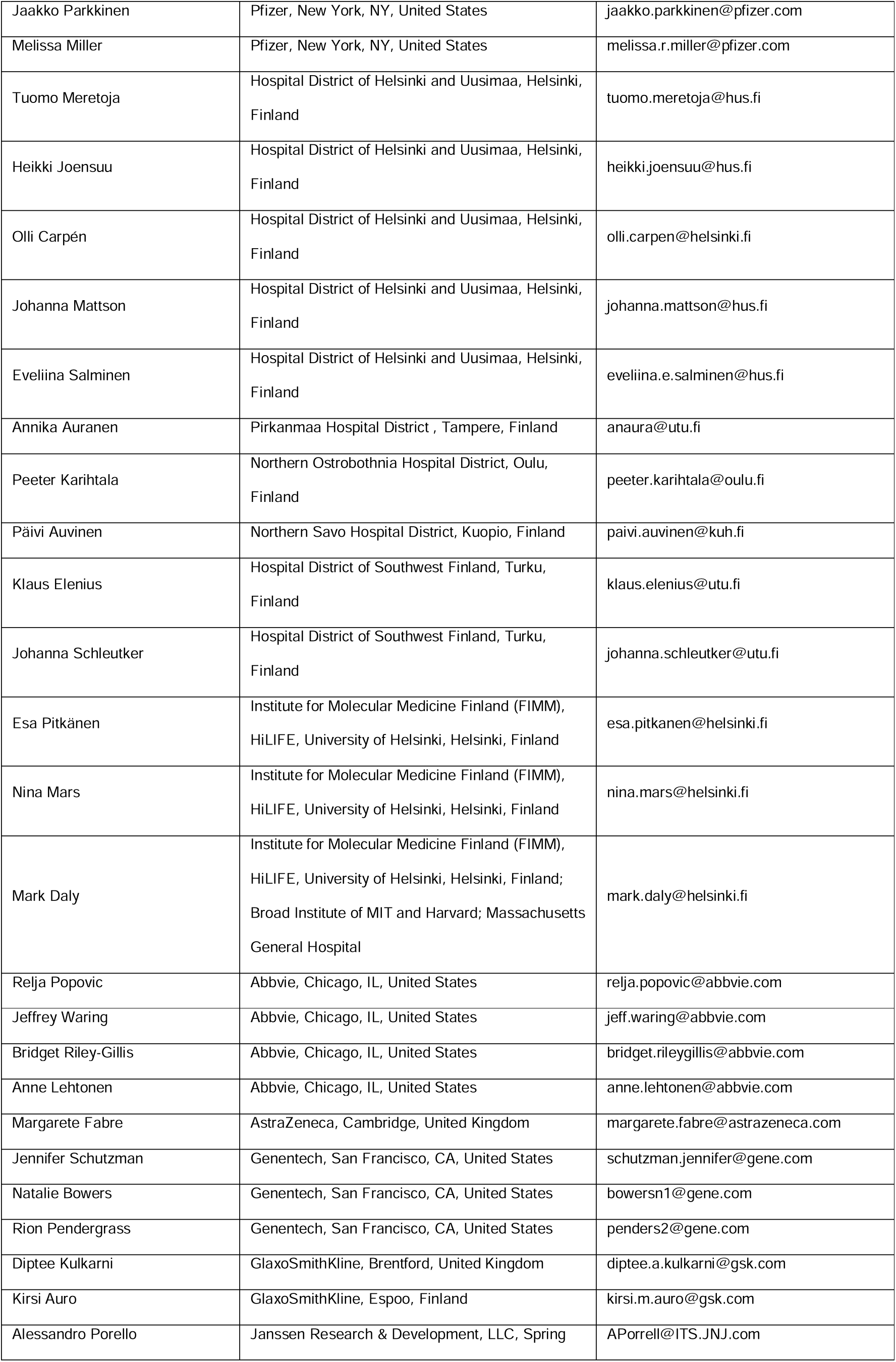
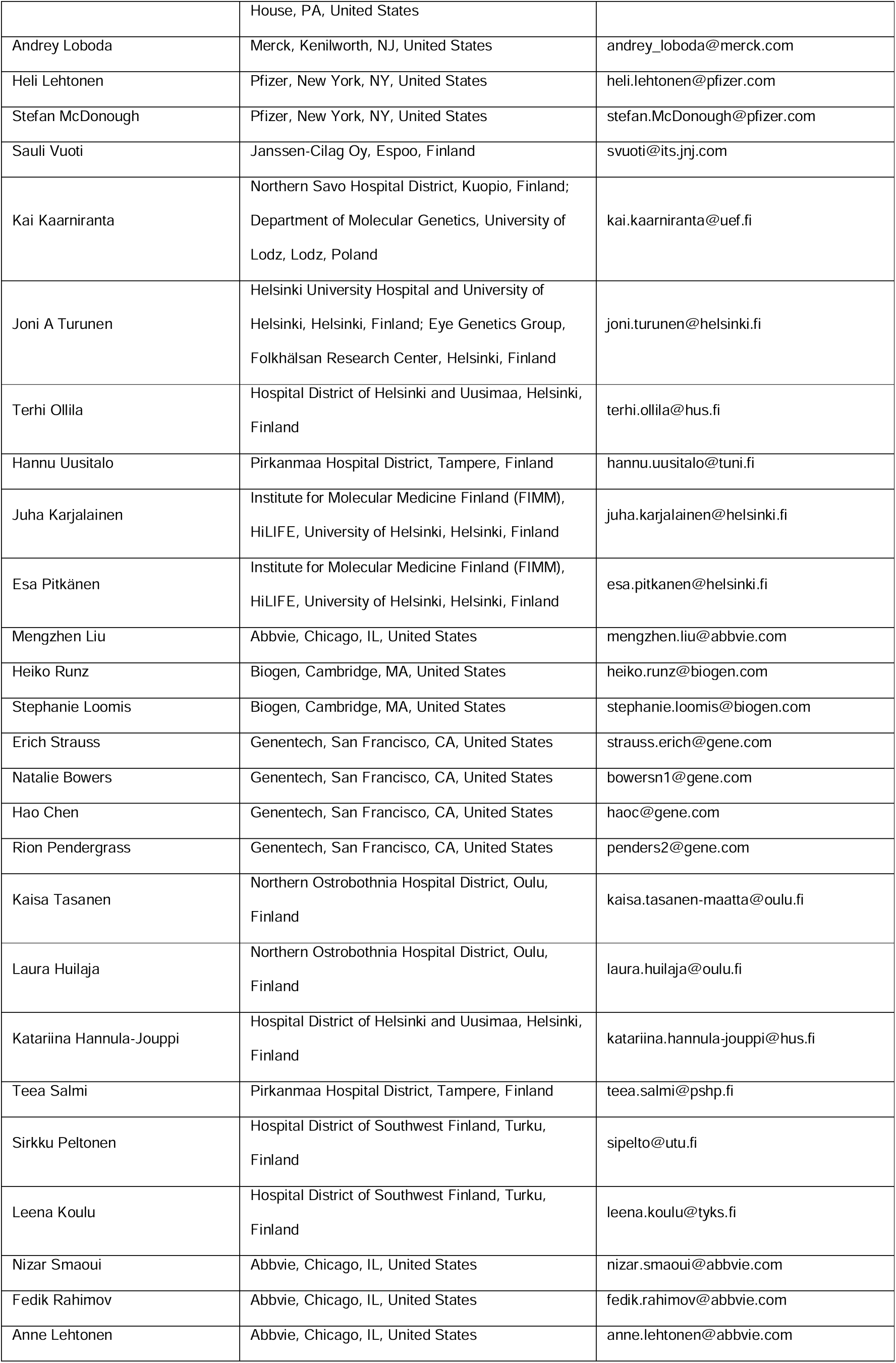
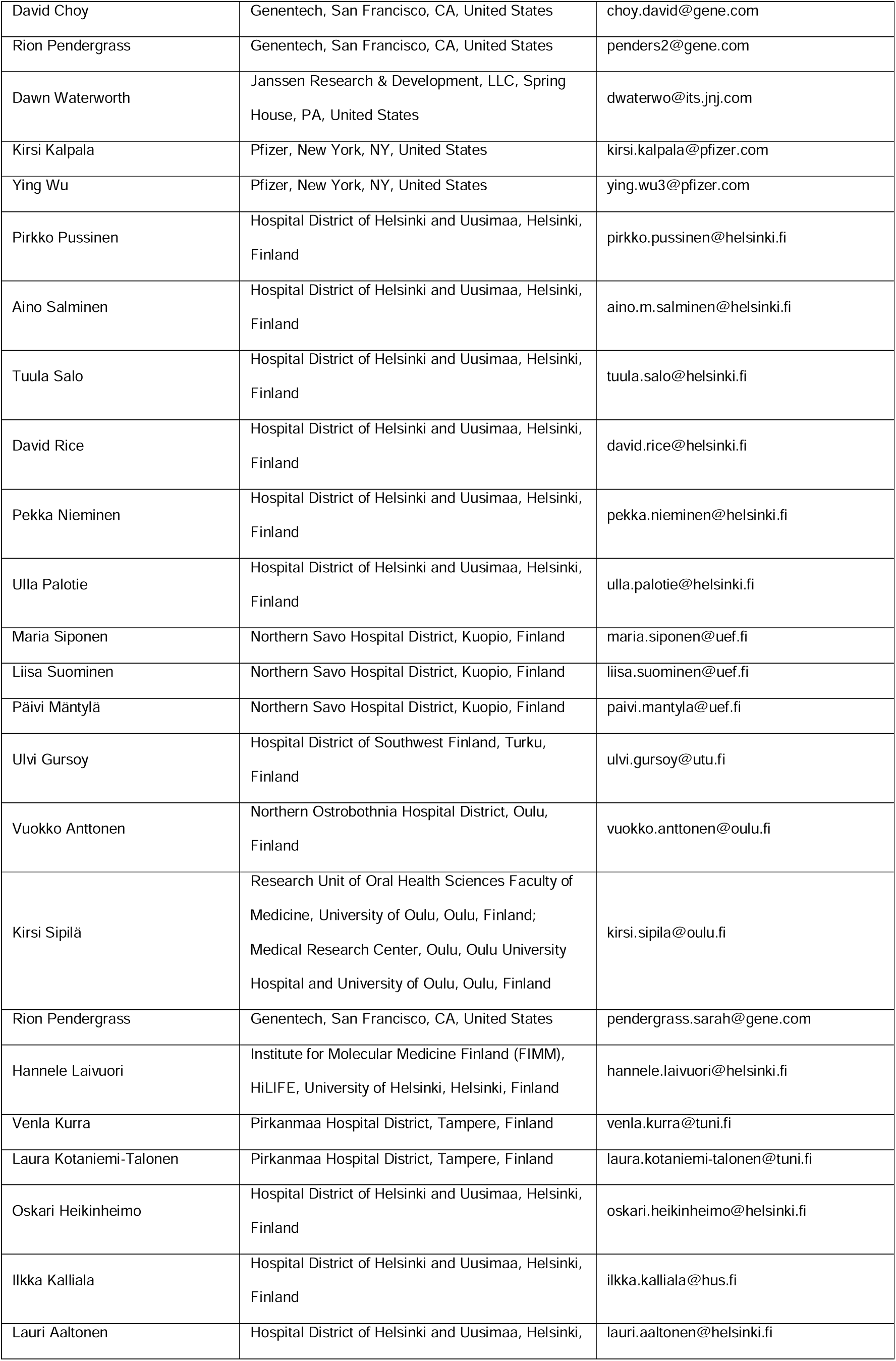
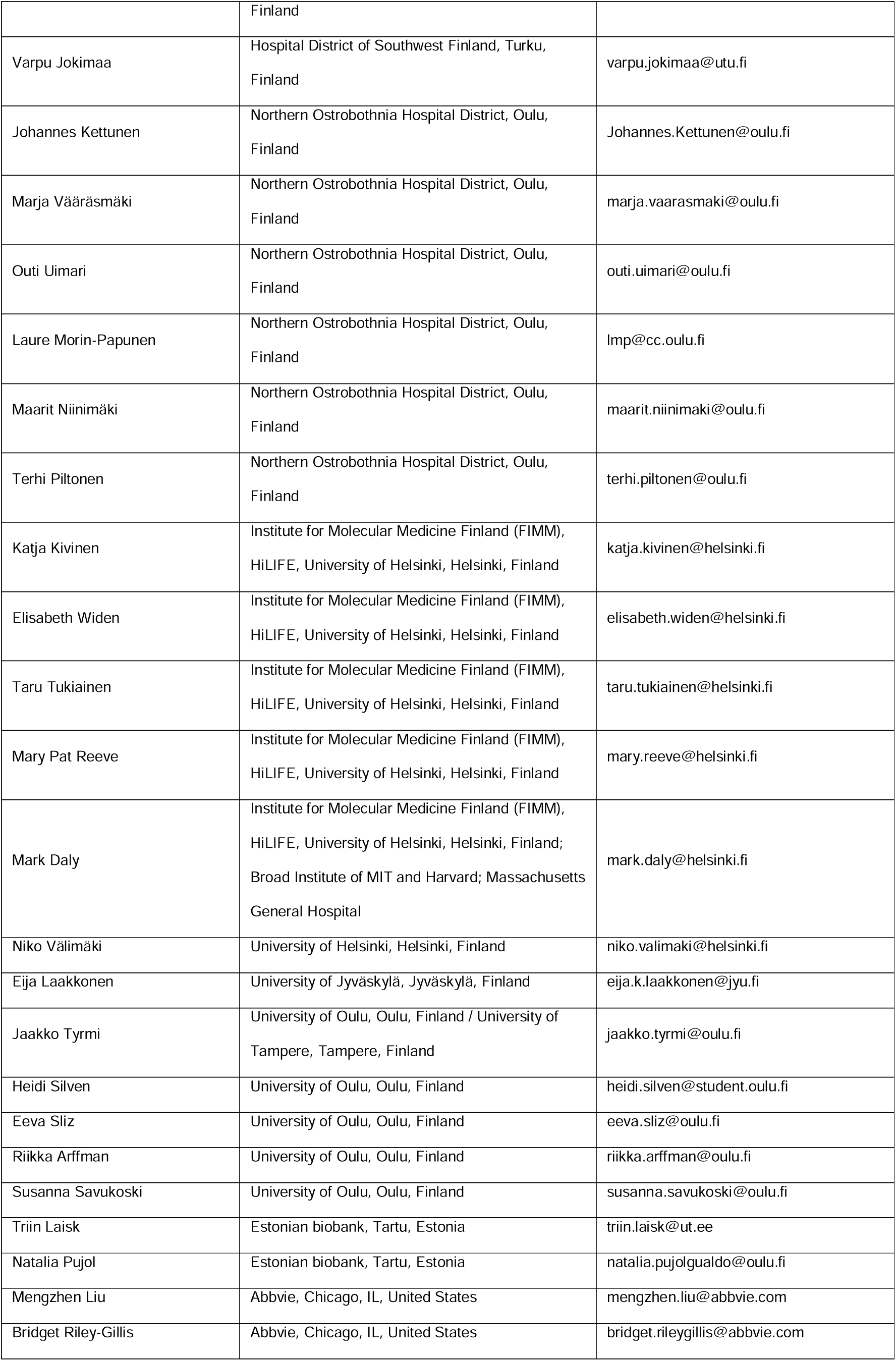
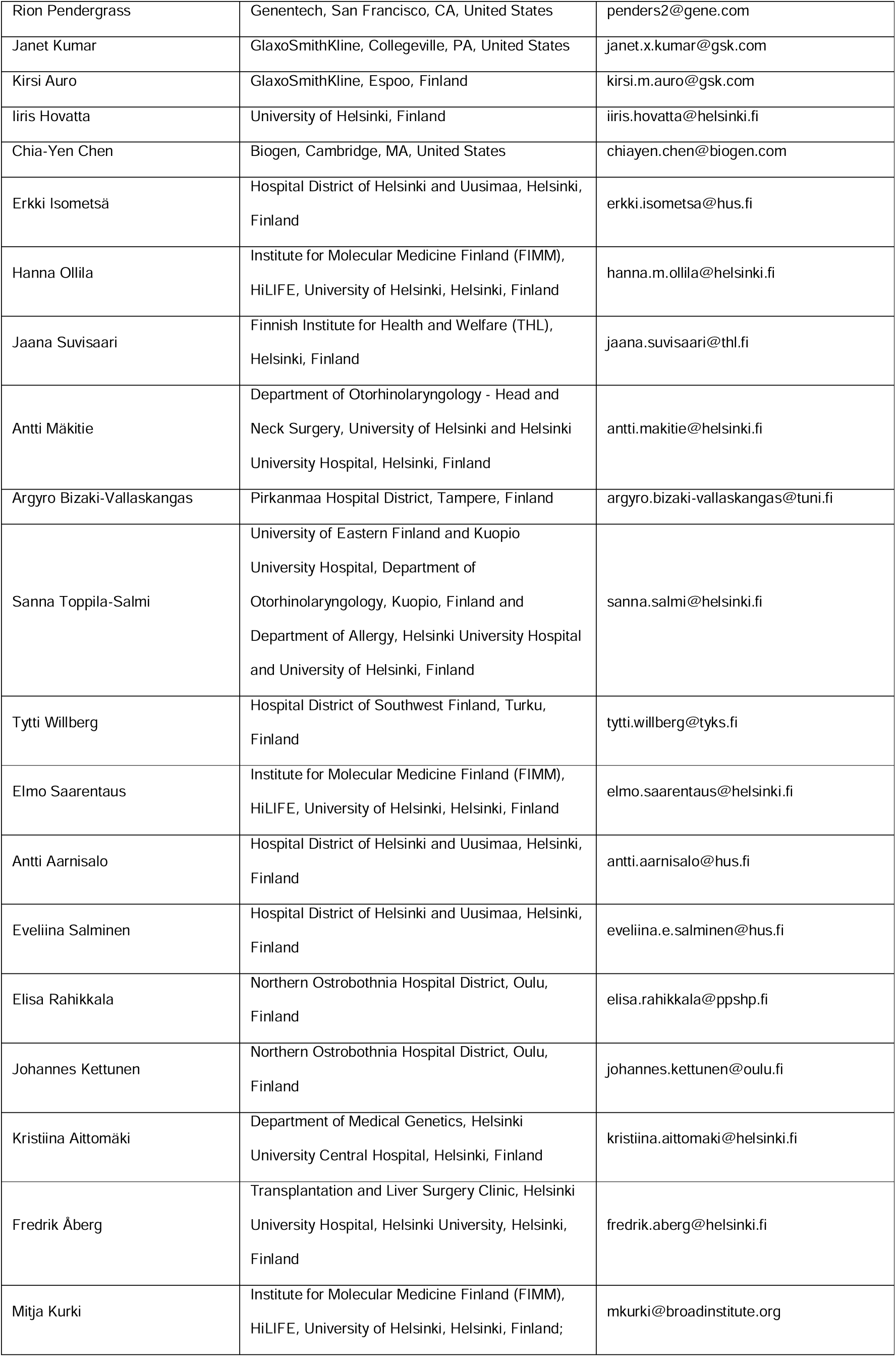
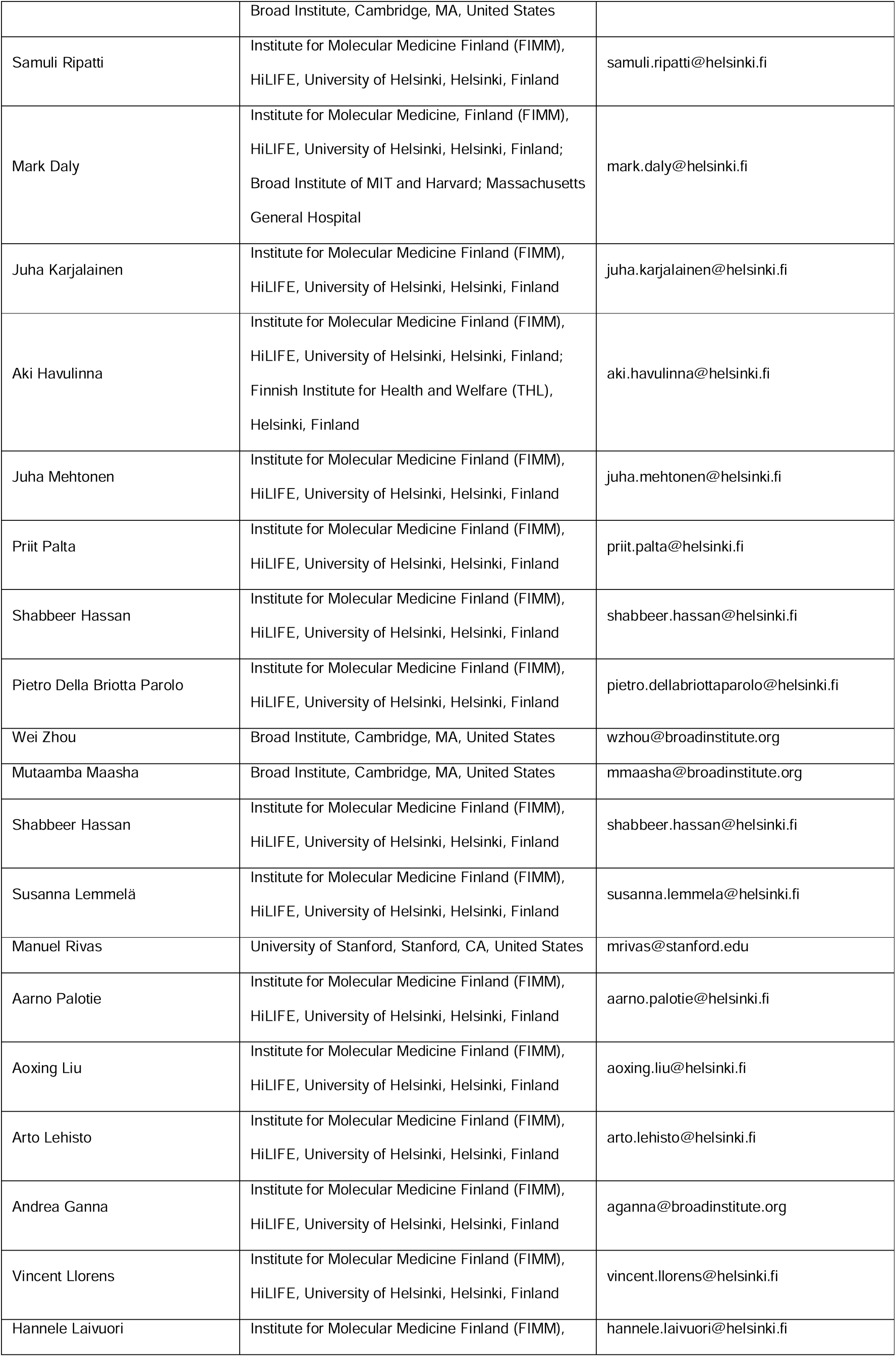
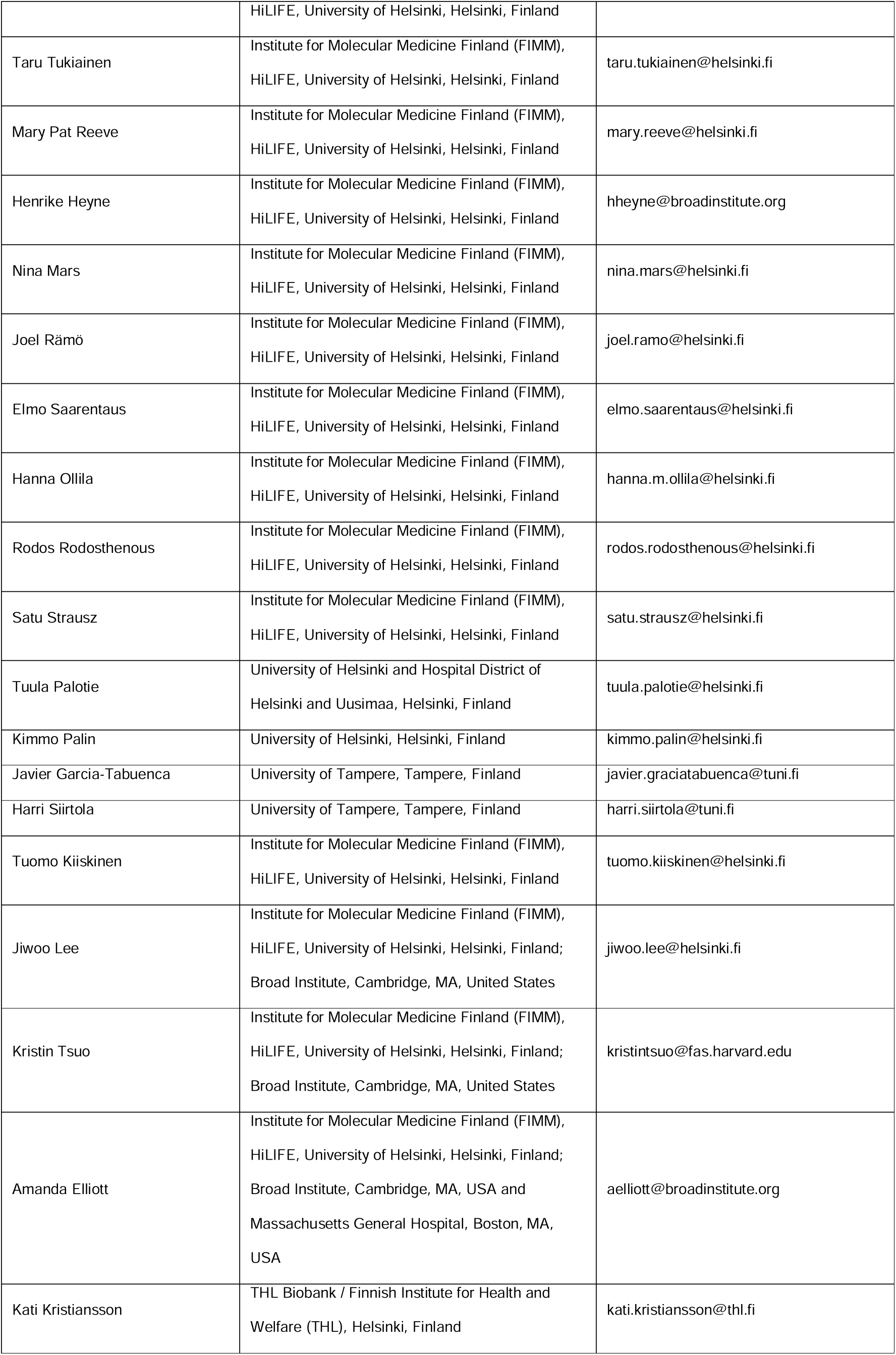
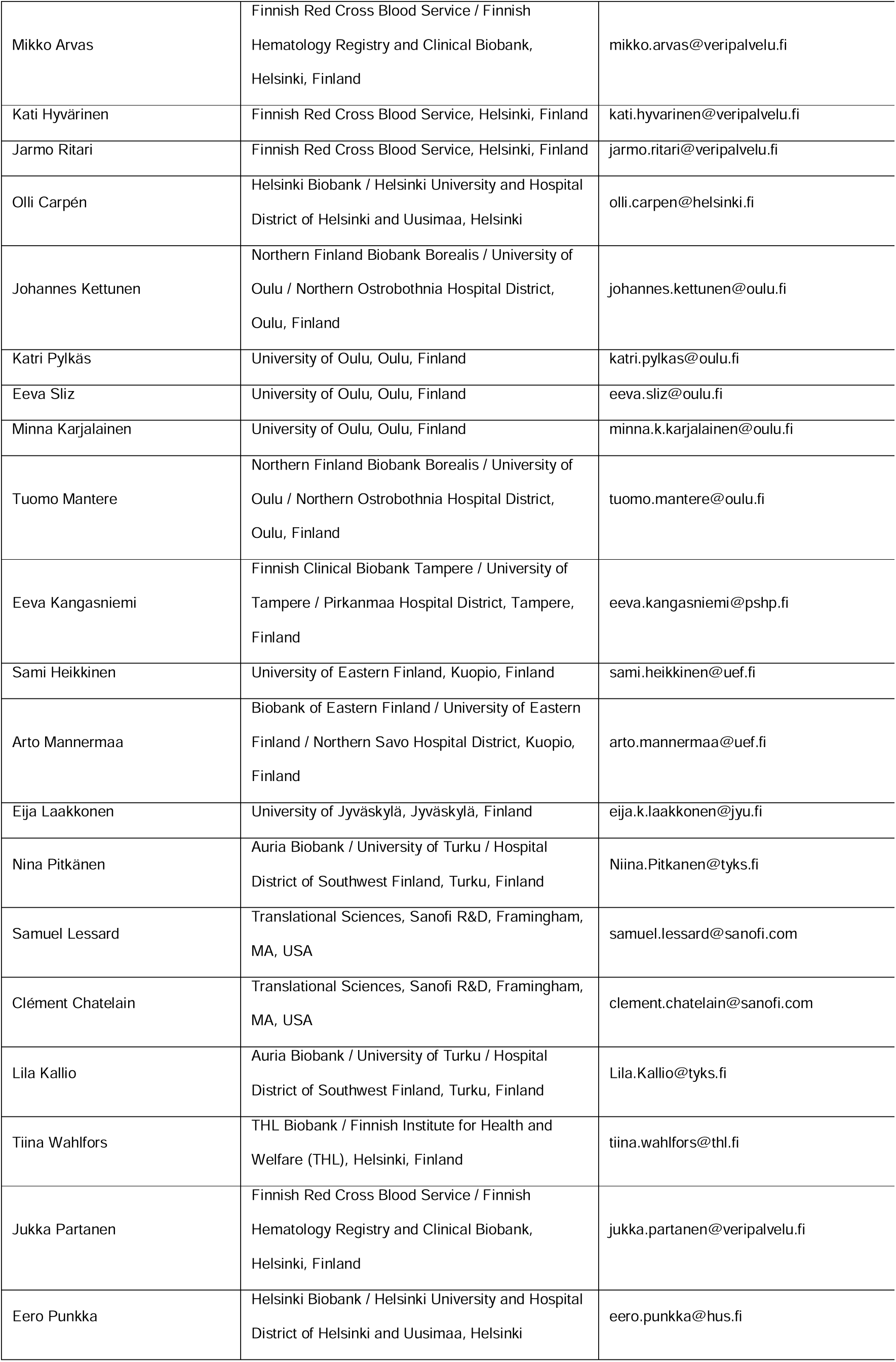
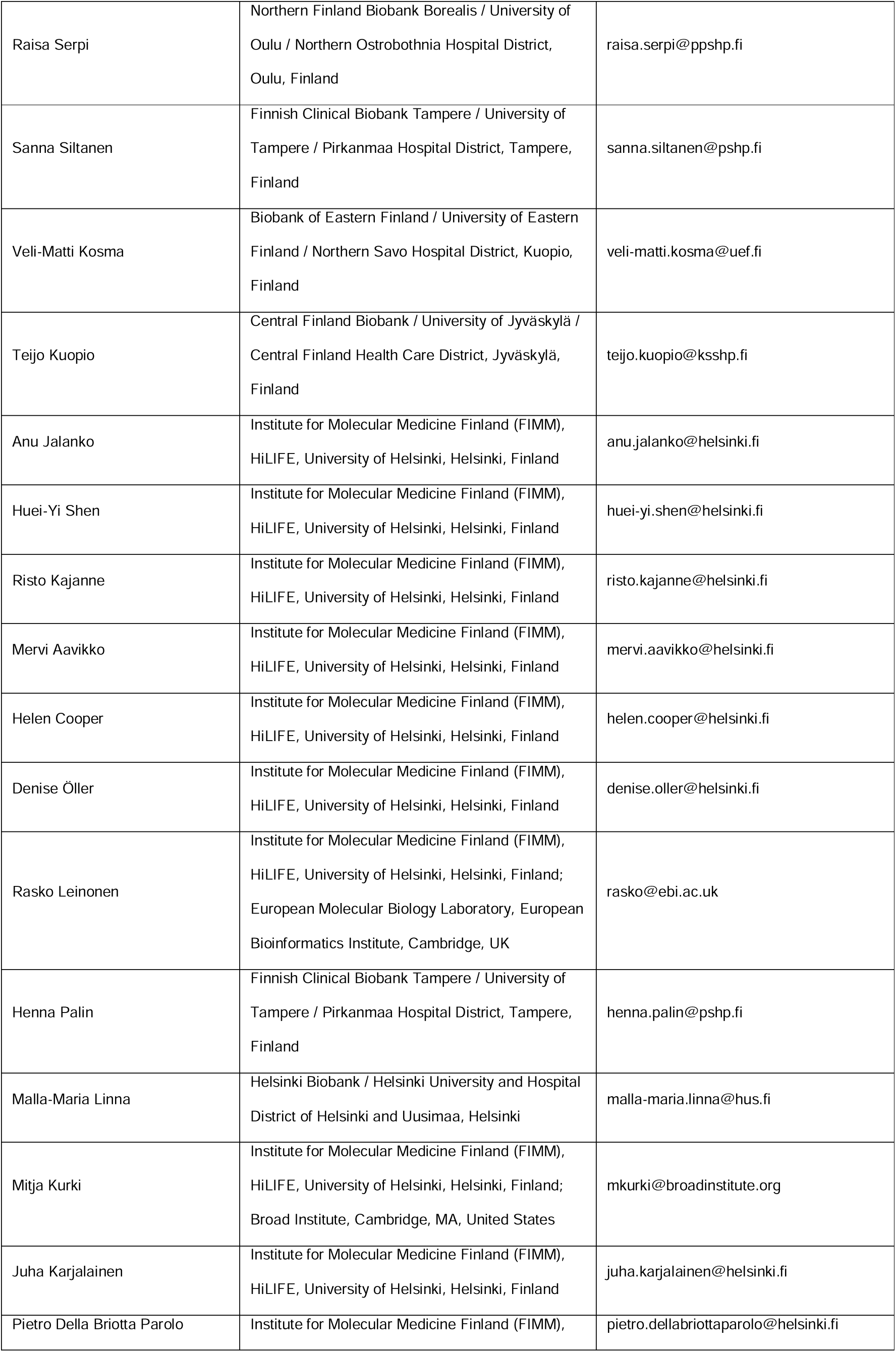
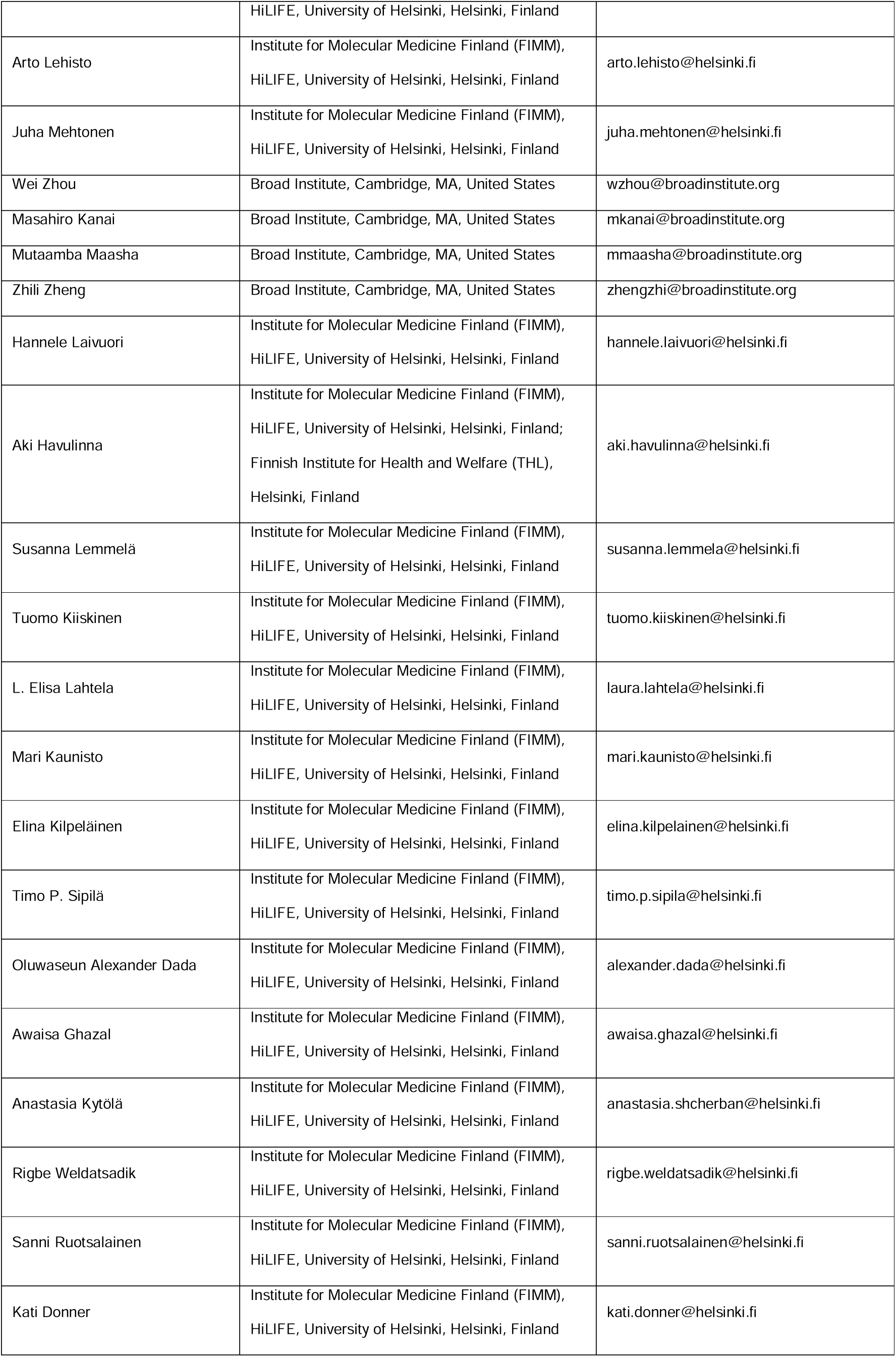
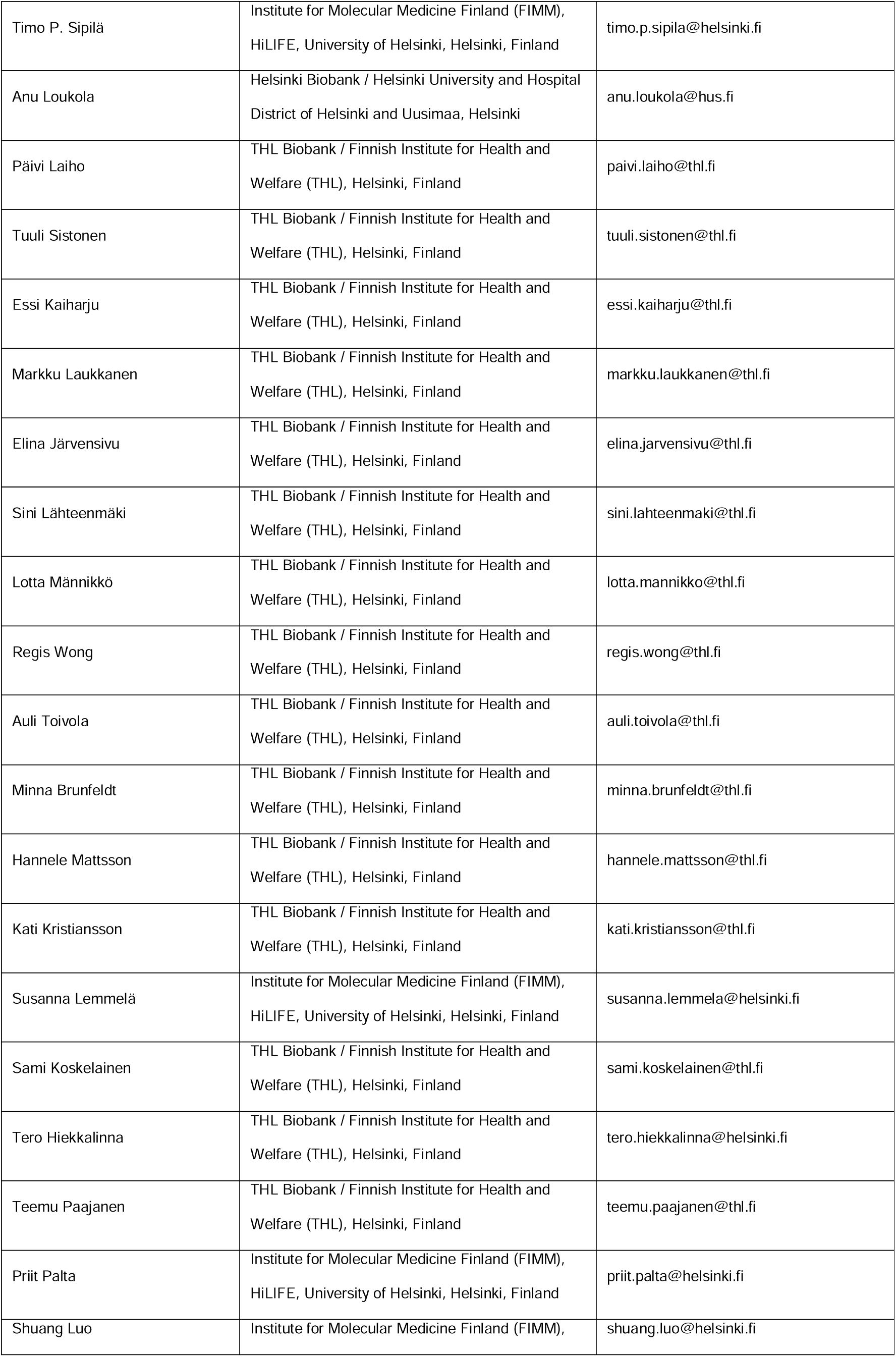
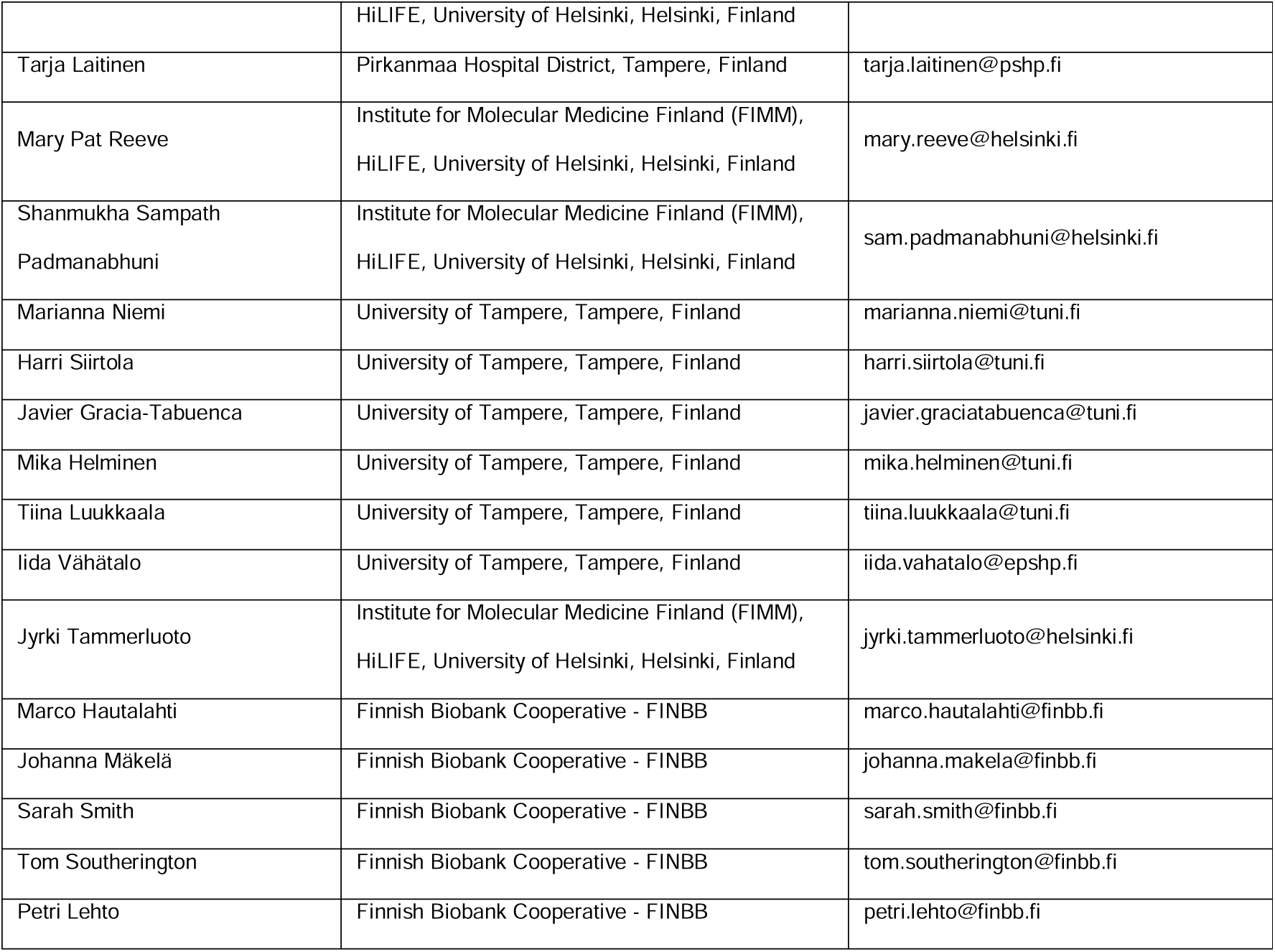
Full list of FinnGen authors.

