## Supplementary Material for "Inherited infertility - mapping loci associated with impaired female reproduction"

### Supplementary figures

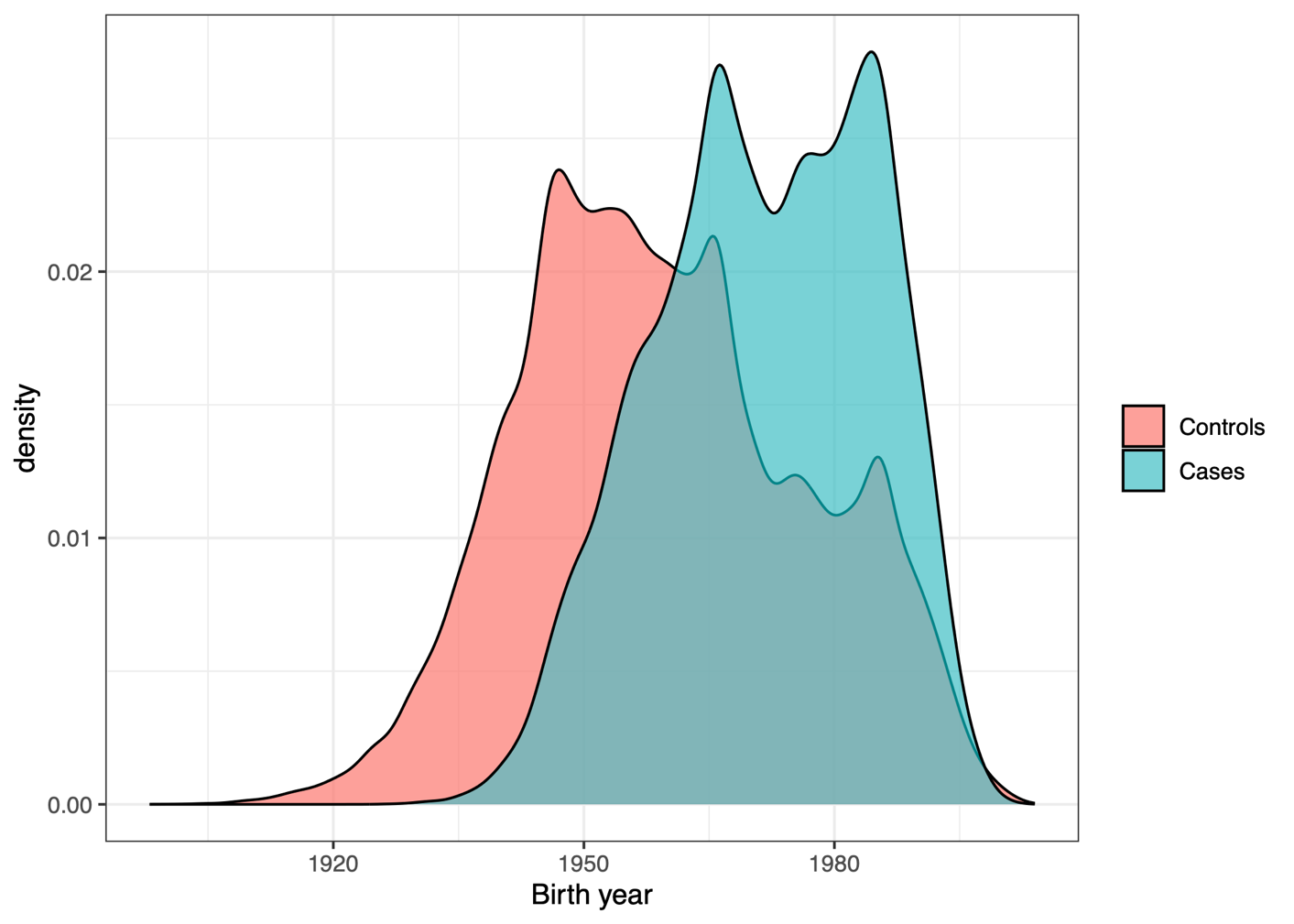

***Supplementary Figure 1:*** *Distribution of birth year for female infertility cases (n = 17,480) and controls (n = 198,989) separately*

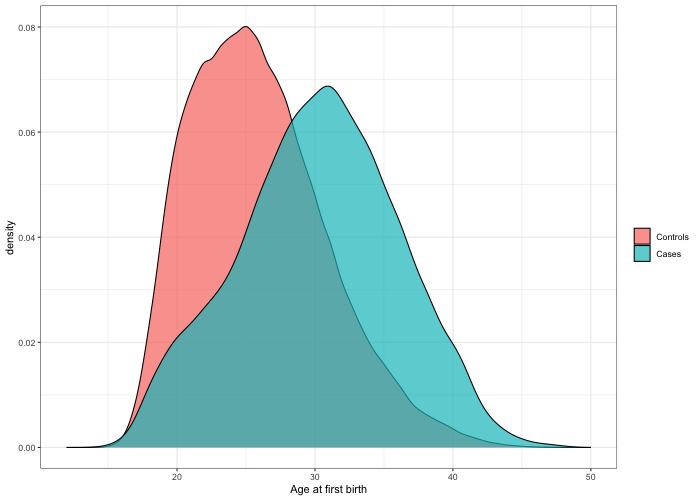

***Supplementary Figure 2:*** *Distribution for age at first birth for female infertility cases and controls separately for those who have given birth at least once (n = 216,469).*

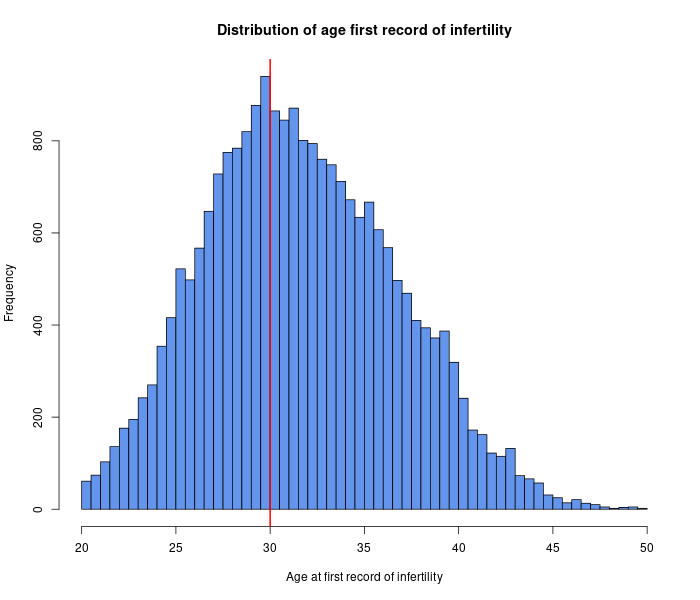

***Supplementary Figure 3:*** *Distribution of age at the first record of female infertility for female infertility cases (n = 22,849)*

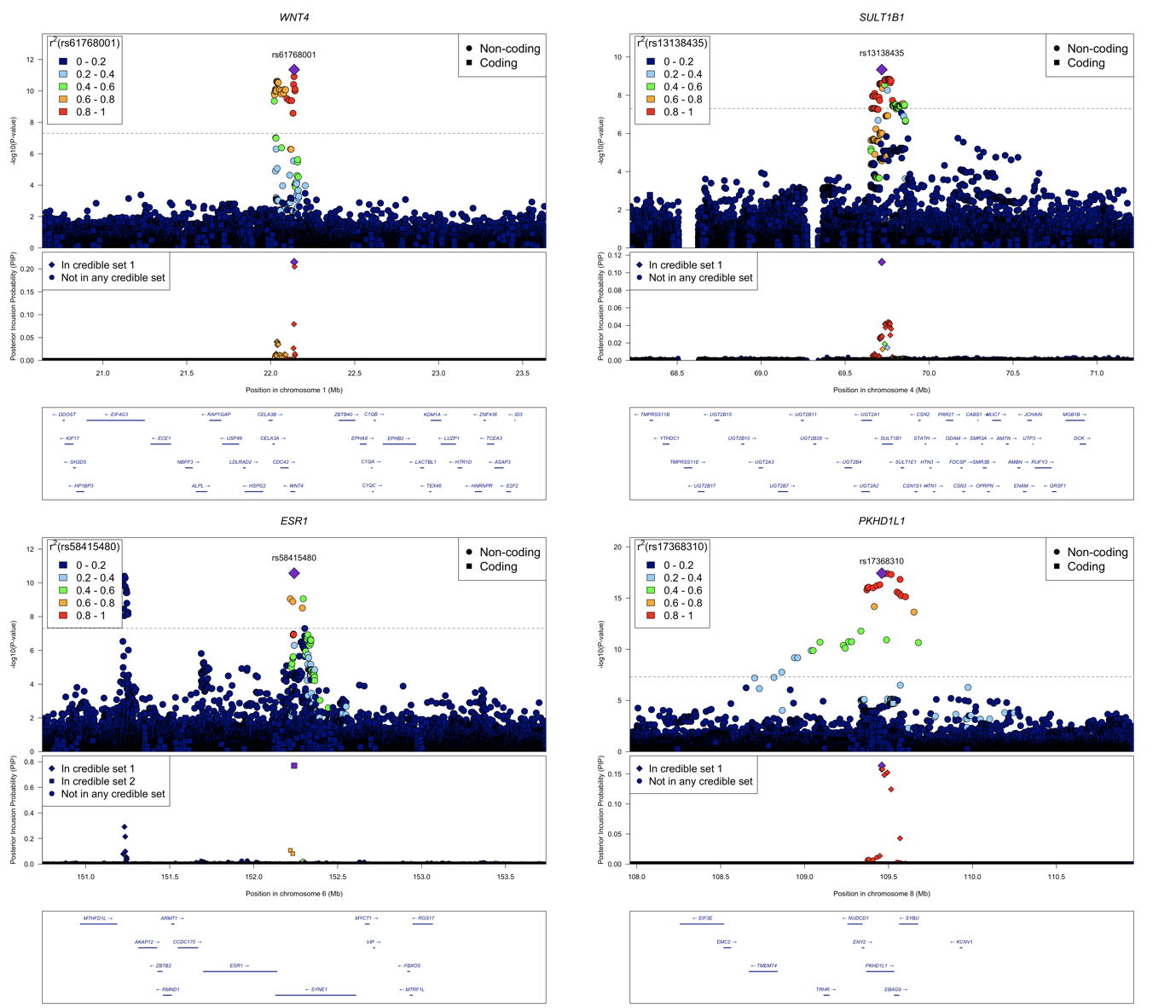

***Supplementary Figure 4:*** *Locuszoomplots from all 4 GWS loci in additive GWAS for female infertility*

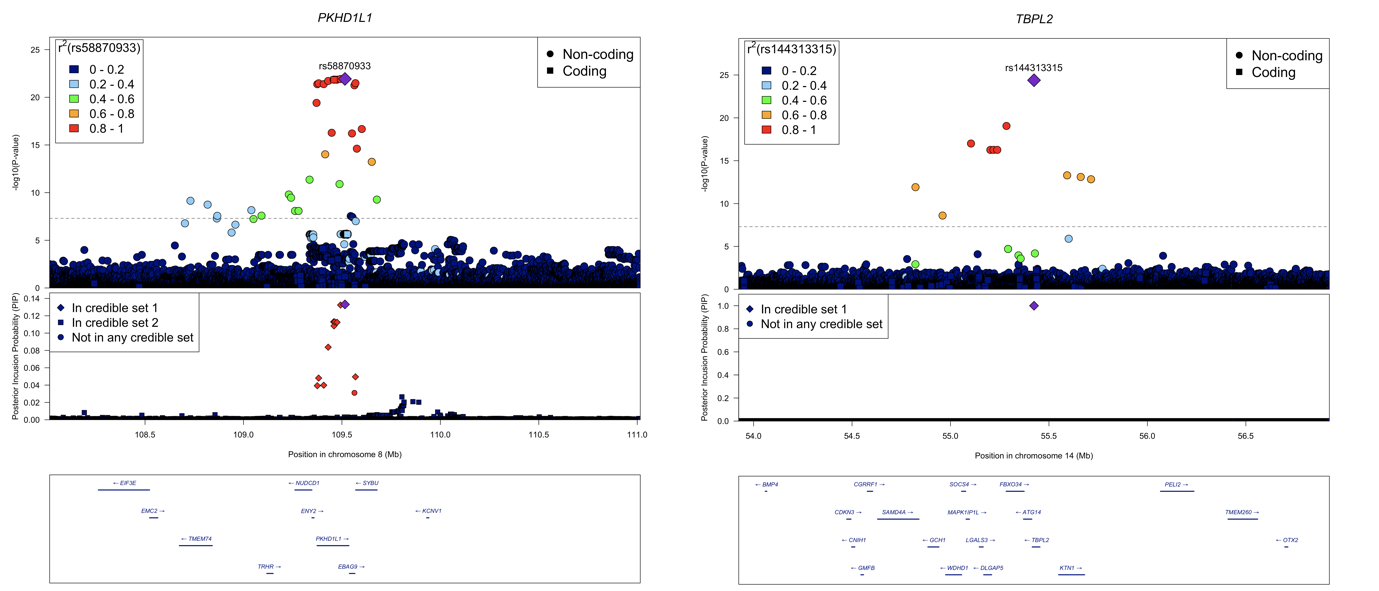

***Supplementary Figure 5:*** *Locuszoomplots from all GWS loci in recessive GWAS for female infertility*

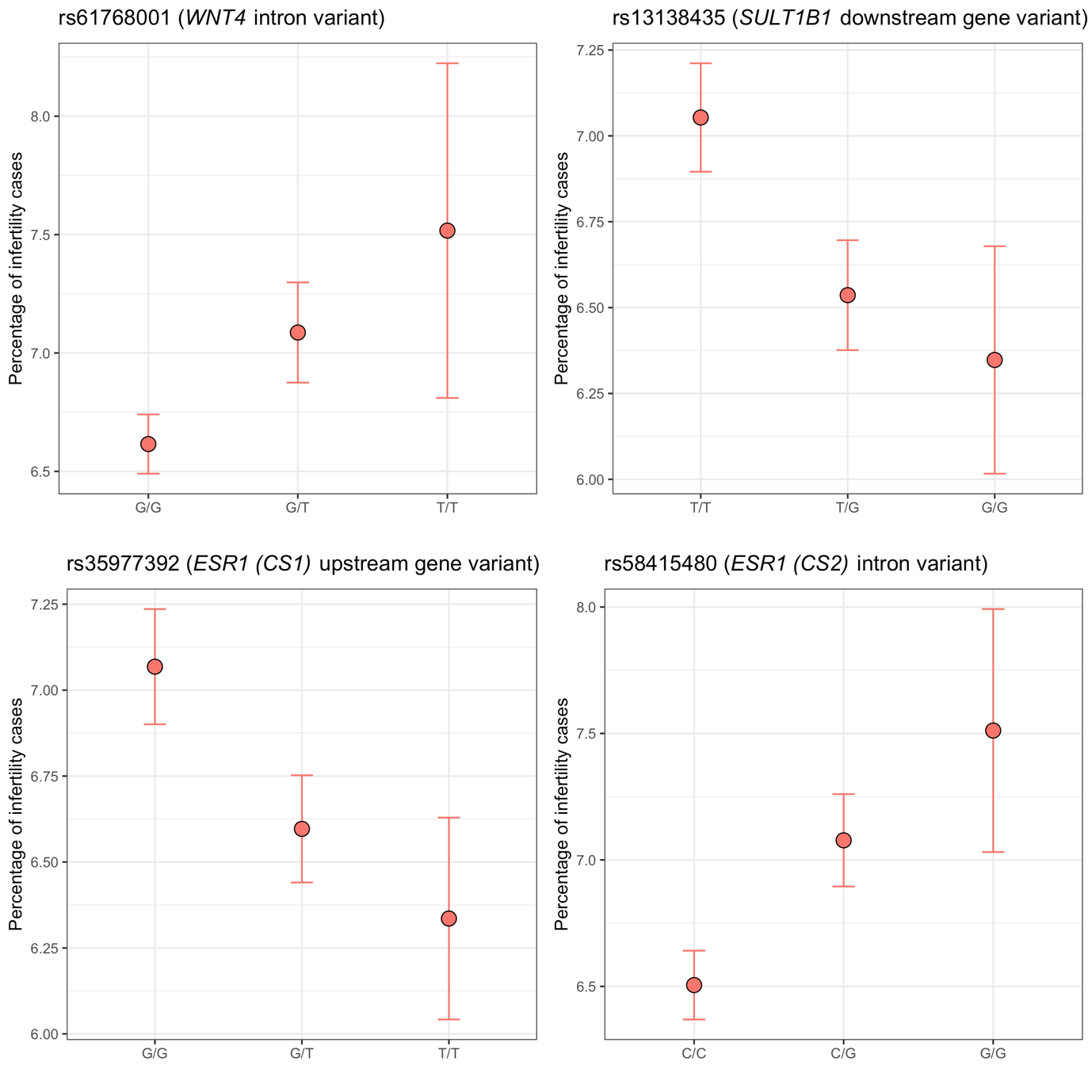

***Supplementary Figure 6:*** *Percentages of infertility cases in genotype groups for all lead variants from the independent signals from the additive scan.*

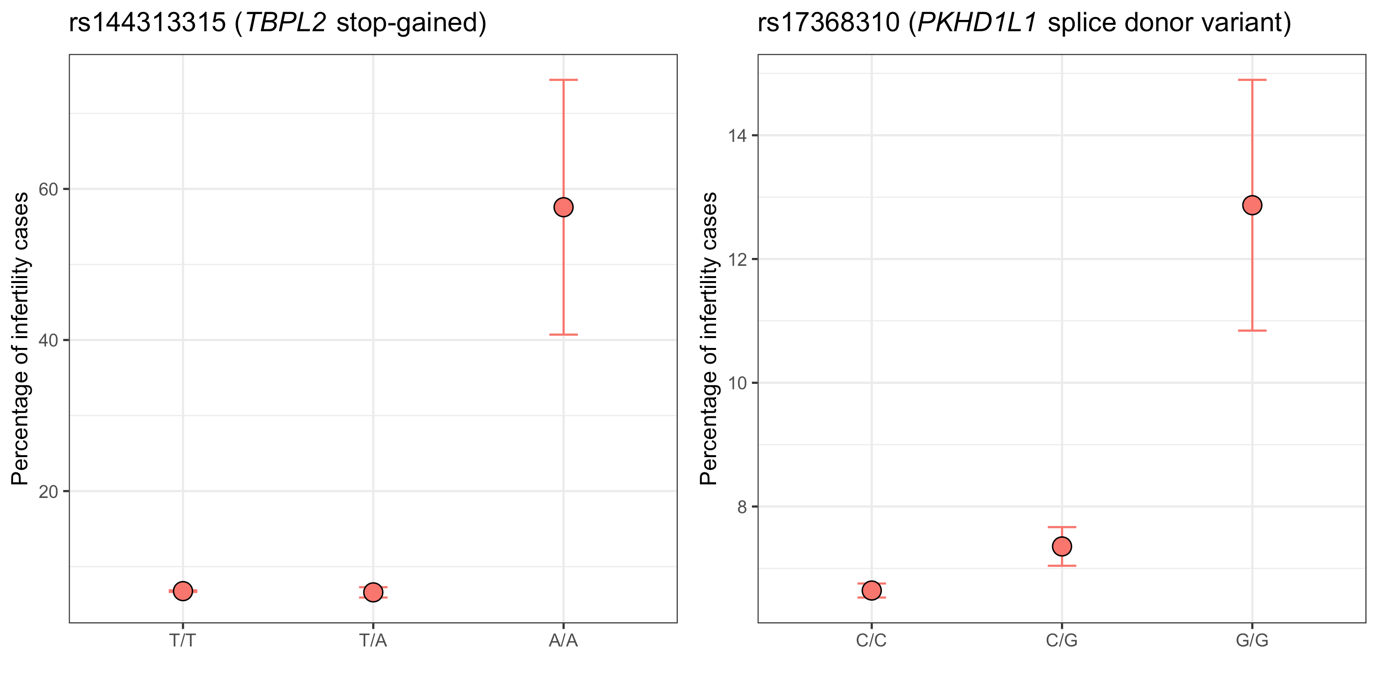

***Supplementary Figure 7:*** *Percentages of infertility cases in genotype groups for all lead variants from the recessive scan.*

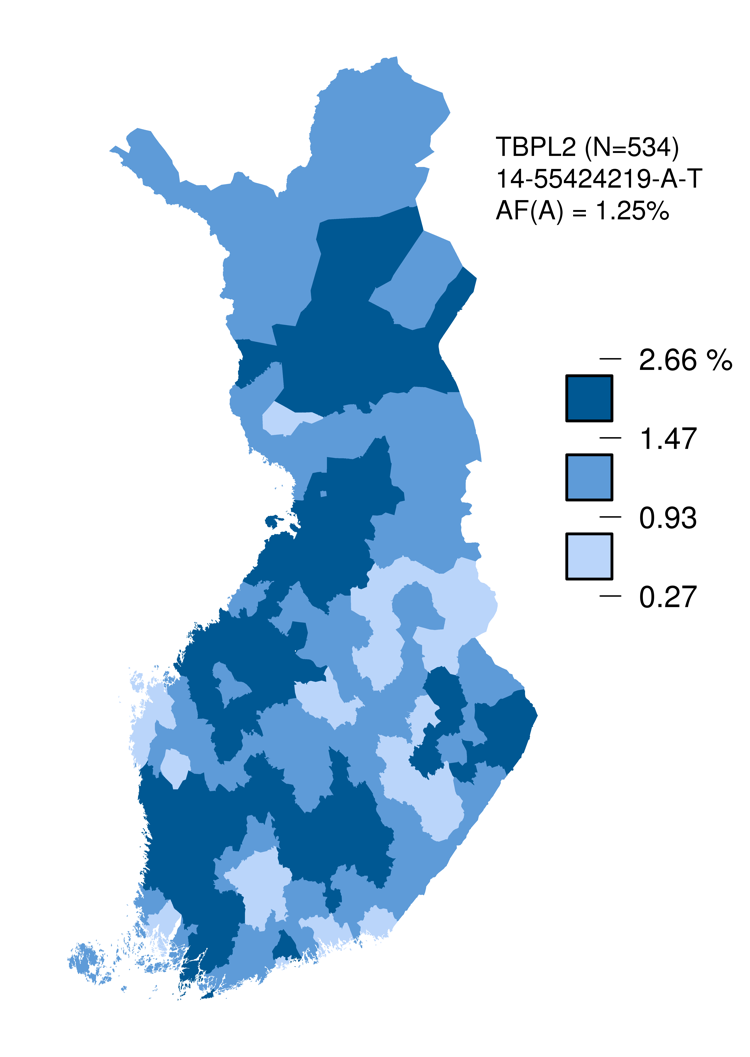

***Supplementary Figure 8:*** *The frequency of allele A of variant rs144313315 in TBPL2 across Finland. Each municipality is colored according to the average allele frequency of 534 individuals closest to the center of the municipality using the color scheme shown next to the map. The genotype data and the municipality of birth was available for 47,950 Finnish individuals from THL biobank (project no. 2019_44). Allele A frequency was 1.25% across the whole data set. The map of Finland with the boundaries of the municipalities was generated by the geoBoundaries R-package (https://www.geoboundaries.org/).*

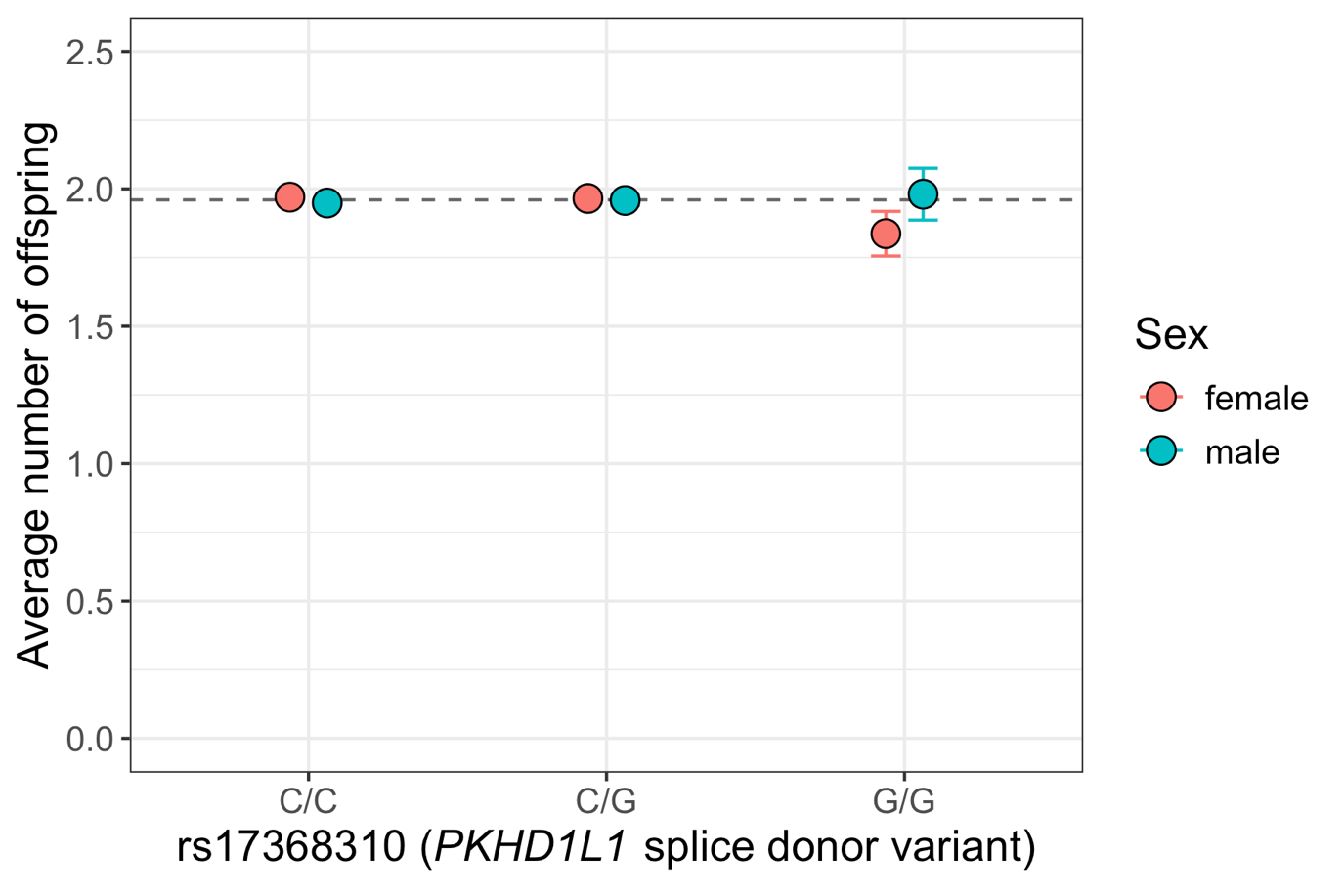

***Supplementary Figure 9****: Average number of offspring for genotype groups for rs17368310 (PKHD1L1 splice donor variant) separately for females and males who have reached age 45. The dashed line represents the overall average number of offspring in FinnGen (=1.96).*

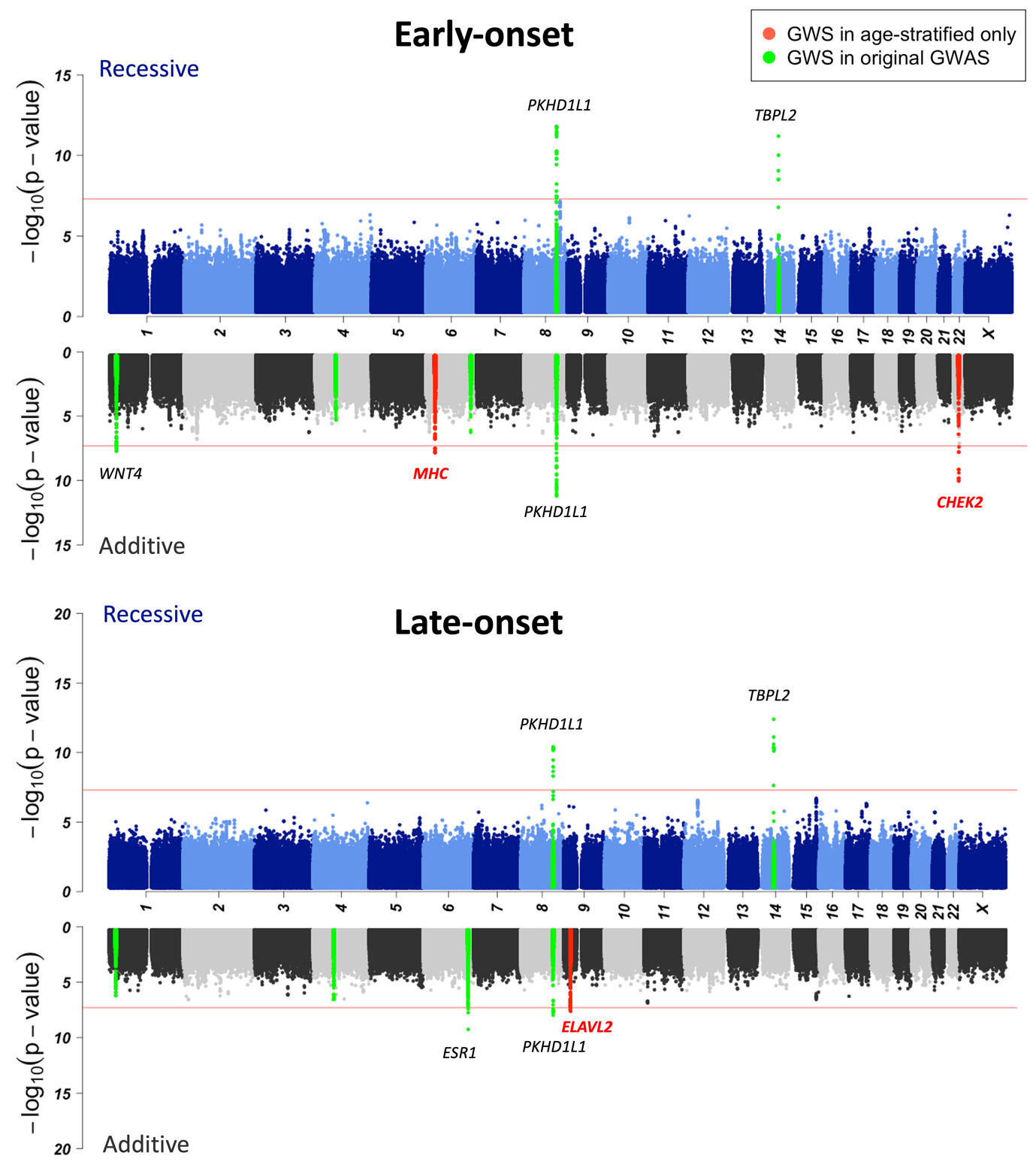

***Supplementary Figure 10:*** *Manhattan plots from age-stratified analyses, for both recessive and additive scan. Genetic loci that are GWS only in the age-stratified analyses are colored as red and genetic loci that were GWS in the original GWAS are colored as green. All GWS loci are labelled with the most severe consequence gene of the lead variant.*

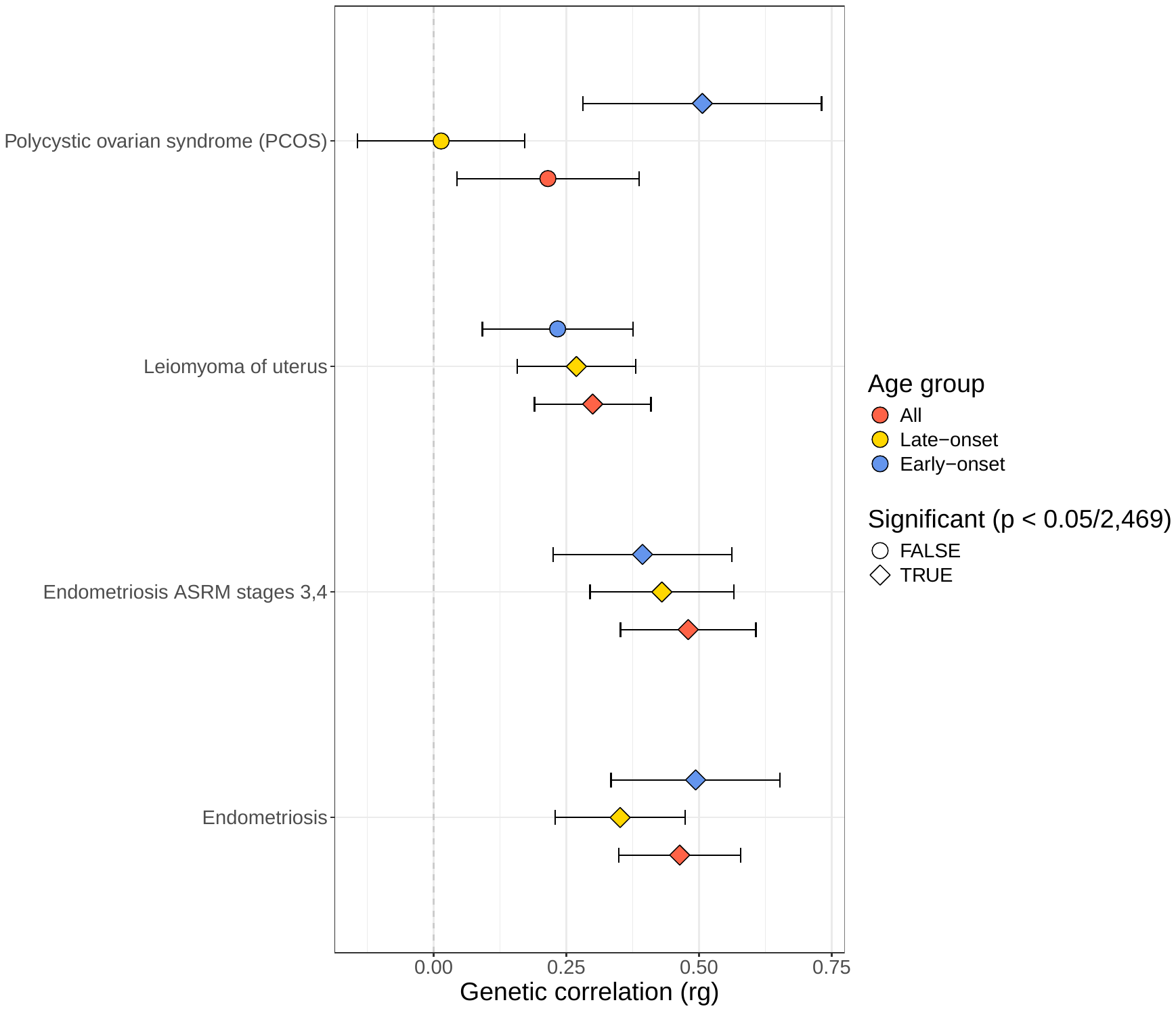

***Supplementary Figure 11:*** *Genetic correlation for 3 female infertility endpoints between 9 female reproduction-related disease endpoints.*

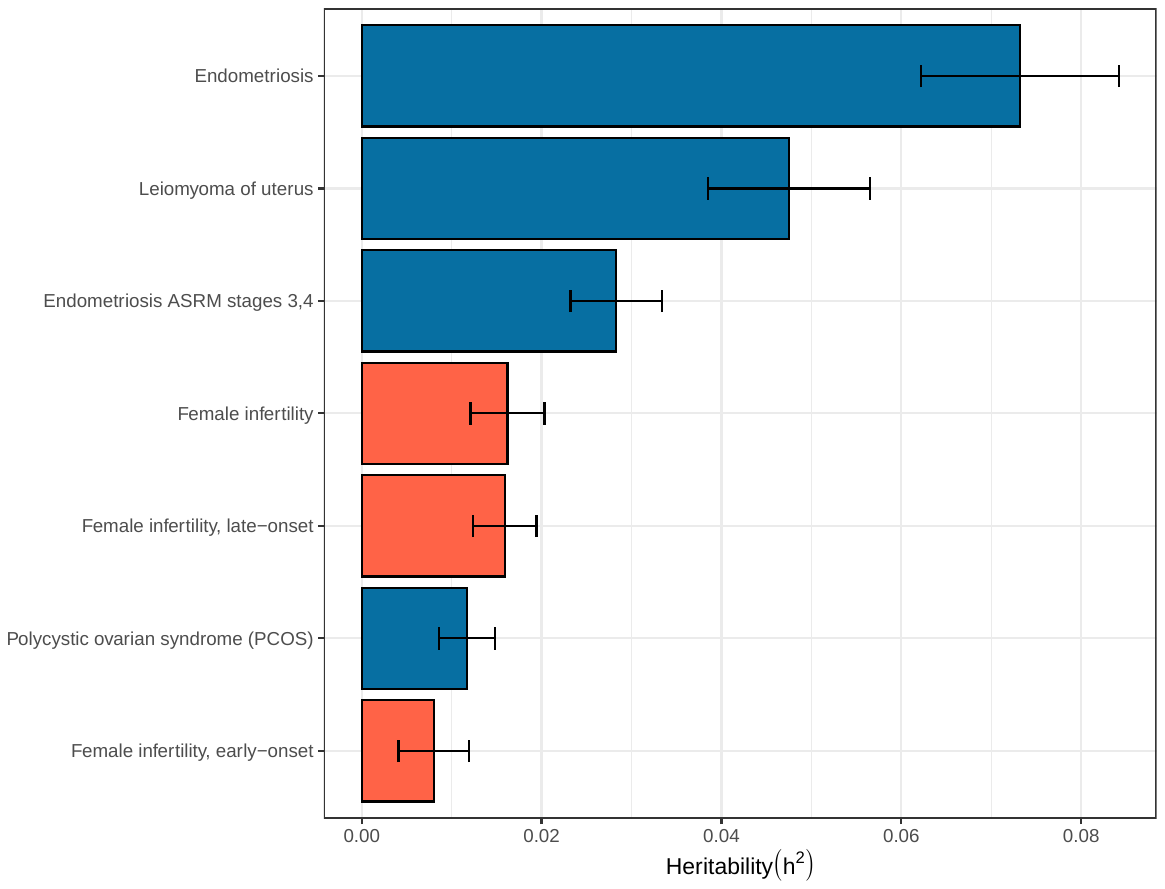

***Supplementary Figure 12:*** *Heritability estimates for 3 female infertility endpoints and 9 female reproduction-related disease endpoints*

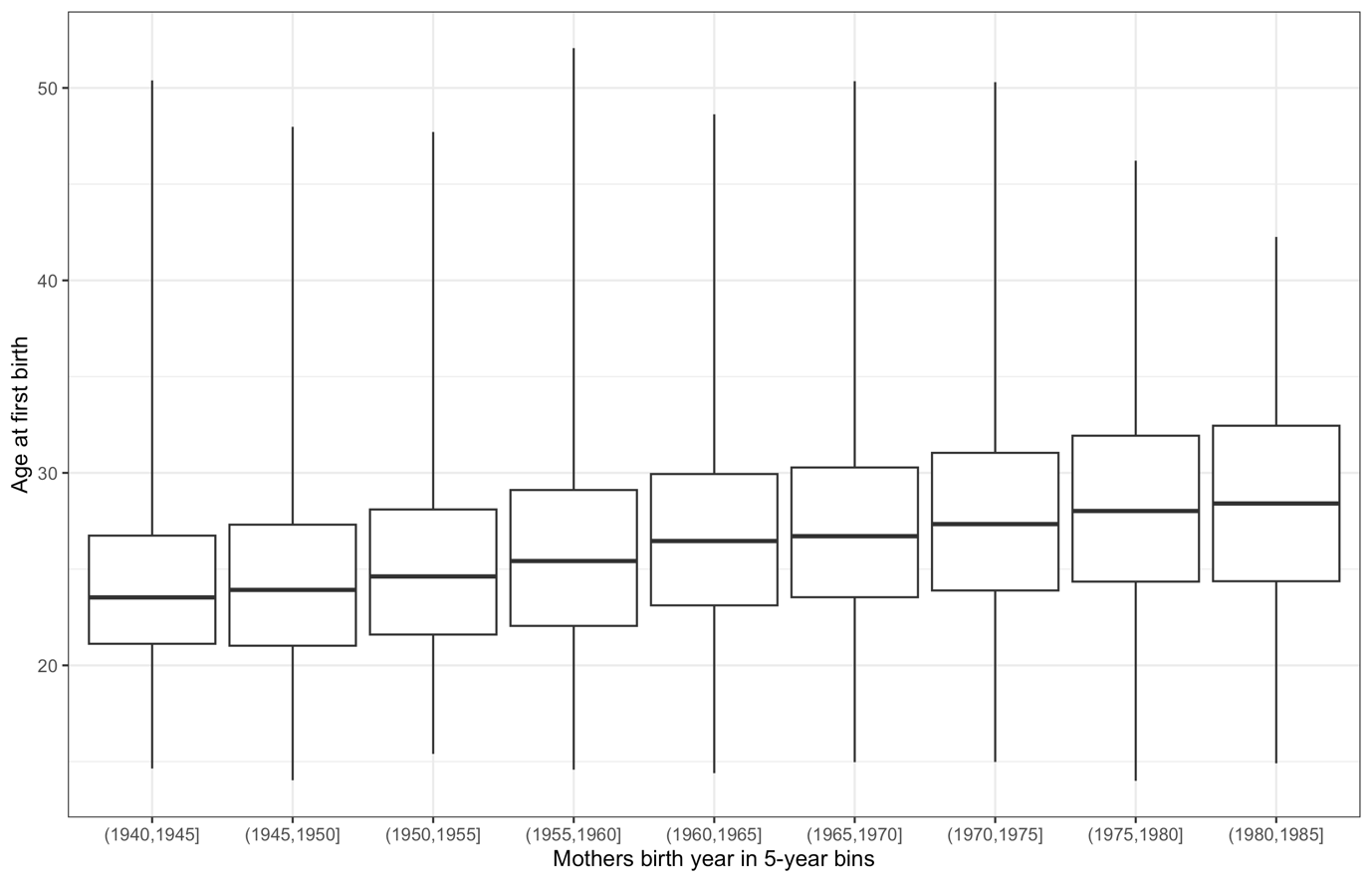

**Supplementary Figure 13:** Distribution of age at first birth for mothers born between 1940-1985.

### Supplementary tables

***Supplementary Table 1:*** *Association results for TBPL2 stop gained variant rs144313315 for all PWS (p < 0.05/2469 = 2.025×10^-05^) phenotypes (n = 5) in either recessive or additive scan*

| **Phenotype** | **Recessive** | | **Additive** | |
| --- | --- | --- | --- | --- |
|  | **OR [95% CI]** | **P-value** | **OR [95% CI]** | **P-value** |
| Female infertility | 11.408 [5.59-23.30] | 2.35×10^-11^ | 1.089 [0.98-1.21] | 0.11 |
| Medical treatment for female infertility | 14.315 [6.17-33.21] | 5.73×10^-10^ | 1.203 [1.04-1.39] | 0.01 |
| Single spontaneous delivery | 0.088 [0.04-0.19] | 1.12×10^-09^ | 0.945 [0.89-0.99] | 0.04 |
| Female infertility, cervigal, vaginal, other or unspecified origin | 10.502 [4.87-22.64] | 1.96×10^-09^ | 1.089 [0.97-1.22] | 0.14 |
| Pregnancy with abortive outcome | 0.074 [0.02-0.22] | 3.60×10^-06^ | 0.982 [0.93-1.04] | 0.52 |
